# The impact of social restrictions during the COVID-19 pandemic on the physical activity levels of older adults: a baseline analysis of the CHARIOT COVID-19 Rapid Response prospective cohort study

**DOI:** 10.1101/2021.01.26.21250520

**Authors:** David Salman, Thomas Beaney, Catherine E. Robb, Celeste A. de Jager Loots, Parthenia Giannakopoulou, Chi Udeh-Momoh, Sara Ahmadi-Abhari, Azeem Majeed, Lefkos T. Middleton, Alison. H. McGregor

## Abstract

**Objectives:** Physical inactivity is more common in older adults, is associated with social isolation and loneliness, and contributes to increased morbidity and mortality. We examined the effect of social restrictions, implemented to reduce transmission of COVID-19 in the UK (lockdown), on physical activity (PA) levels of older adults, and the demographic, lifestyle and social predictors of this change.

**Design:** Baseline analysis of a survey-based prospective cohort study

**Setting:** Adults enrolled in the Cognitive Health in Ageing Register for Investigational and Observational Trials (CHARIOT) cohort from GP practices in North West London were invited to participate from April to July 2020.

**Participants:** 6,219 cognitively healthy adults aged 50 to 92 years completed the survey.

**Main outcome measures:** Self-reported PA before and after lockdown, as measured by Metabolic Equivalent of Task (MET) minutes. Associations of PA with demographic, lifestyle and social factors, mood and frailty.

**Results:** Mean PA was significantly lower following lockdown, from 3,519 MET minutes/week to 3,185 MET minutes/week (p<0.001). After adjustment for confounders and pre-lockdown PA, lower levels of PA after lockdown were found in those who were over 85 years old (640 [95% CI: 246 to 1034] MET minutes/week less); were divorced or single (240 [95% CI: 120 to 360] MET minutes/week less); living alone (277 [95% CI: 152 to 402] MET minutes/week less); reported feeling lonely often (306 [95% CI: 60 to 552] MET minutes/week less); and showed symptoms of depression (1007 [95% CI: 1401 to 612] MET minutes/week less) compared to those aged 50-64 years, married, co-habiting, and not reporting loneliness or depression, respectively.

**Conclusions and Implications:** Markers of social isolation, loneliness and depression were associated with lower PA following lockdown in the UK. Interventions to improve PA in older adults should take account of social and community factors, and targeted strategies to increase physical activity in socially isolated, lonely and depressed older adults should be considered.

## 1.0 Background and Rationale

Physical inactivity (PA) adversely affects older adults, with 60-70% of those aged over 75 years not sufficiently active for good health^1,2^ as defined by meeting World Health Organization (WHO)^3^ and UK^4^ guidelines. From March until June 2020 in the UK, a national ‘lockdown’ was implemented to reduce exposure to, and transmission of, COVID-19. Although applied to the whole population, adults aged over 70 years and those with underlying health conditions at higher risk of severe COVID-19 disease were asked to follow more stringent social distancing measures. These included remaining at home where possible; avoiding social mixing in the community; avoiding physically interacting with friends and family; and avoiding public transport.^5^

Social isolation and loneliness in older adults, possibly exacerbated during lockdowns,^6^ is associated with increases in morbidity and mortality, and also with increases in physical inactivity and sedentary time, as shown from subjective self-reporting and from accelerometer data.^7,8^ Physical inactivity may therefore have a role in mediating the increased morbidity and mortality associated with social isolation.^9^ We set up the CHARIOT COVID-19 Rapid Response study (CCRR) in April 2020 to monitor symptoms and the impact of the COVID-19 pandemic on various health and lifestyle factors, by repeat questionnaire survey of the Cognitive Health in Ageing Register for Investigational and Observational Trials (CHARIOT) members.

We hypothesised that imposed social restrictions would negatively impact on PA levels of older adults, and that change in PA after lockdown would be modified by certain demographic, lifestyle and social factors, with a focus on markers of social isolation and perceived loneliness. An awareness of the extent of, and predictors for, change in PA levels will aid our understanding of the impact of social isolation on the health of older adults, both with respect to pandemic-related lockdowns and social isolation itself.

## 2.0 Methods

### 2.1 CCRR survey

Study participants were recruited from the CHARIOT register, a cohort of over 40,000 cognitively healthy adult volunteers aged over 50 years, recruited from 172 GP surgeries across West and North London as part of a collaboration between regional GP practices and the School of Public Health, at Imperial College London. This ongoing prospective cohort study was initiated in April 2020 with repeated questionnaire surveys conducted every six weeks. The CCRR baseline survey consists of questions related to basic demographics, diet, alcohol and smoking status, symptoms of COVID-19, functional activities, physical activity, sleep, frailty and mental health (supplementary file 1). For physical activity, the International Physical Activity Questionnaire (IPAQ) short-form was used,^10^ asking respondents to document their weekly vigorous and moderate activity, walking and sitting time from the week prior to completing the survey; and for the week prior to implementation of social restriction measures. For assessing frailty, the 5-point FRAIL scale,^11,12^ and for assessing mental health symptoms, the Hospital Anxiety and Depression (HADS) scale,^13^ were used. A question on loneliness was used from the Imperial College Sleep Quality questionnaire; in turn adapted from the Pittsburgh Sleep Quality Index^14^ and Centre for Epidemiologic Studies of Depression Scale^15^, for work-free periods.

The survey was sent to 15,000 CHARIOT participants via email, with a subsequent 25,000 contacted by post. 7,320 participants responded and completed the survey. Of these respondents, 6,219 completed IPAQ data both before and after introduction of lockdown measures and were included in the final analysis. Data used in the present analysis were completed between 30^th^ April and the 22^nd^ July 2020.

### 2.2 Statistical analysis

All analyses were conducted using Stata version 16.1 (StataCorp 2019) and R.^16,17^ Body Mass Index (BMI) was calculated as weight in kilograms divided by the square of height in metres and categorised according to standard WHO criteria. IPAQ data were cleaned according to the IPAQ data cleaning protocol,^18^ and the Metabolic Equivalent of Task (MET) minutes per week, calculated for each activity and total activity (supplementary file 2). Paired t-tests were used to compare the distributions of mean PA levels pre- and post-lockdown.

Measures of association with explanatory variables were explored in univariable linear regression models for two outcomes: i) overall weekly MET minutes after introduction of lockdown and ii) the difference in overall weekly MET minutes before versus after the introduction of lockdown. Multivariable models were constructed for the outcome of MET minutes after lockdown, adjusting *a priori* each explanatory variable in turn for age, sex and ethnicity. Month of survey completion was also included in the model to account for seasonal changes, and the finding that physical activity after lockdown varied by month (supplementary file 2: figure 2). Weekly MET minutes before lockdown was also included in the model given its strong association with activity levels after lockdown, which remained significantly associated in all models. Denominators for each model vary according to the levels of missingness in variables included in the models, which was low for most variables, except for BMI (unrecorded in 51.4% of participants).

A causal diagram was constructed using DAGitty^19^ (supplementary file 2: figure 4) to aid adjustment for confounders in order to separate the overall causal effects of marital status, loneliness and living alone on physical activity. Additional multivariable models were then constructed based on the causal diagram for loneliness, adjusting for age, sex, ethnicity, household status, marital status, shielding status and frailty category. No further adjustment was necessary for marital status or household status. Residuals were plotted against fitted values to assess for outlying points and heteroskedasticity; and plots of Cook’s distance and leverage against fitted values were examined to detect the presence of influential points.

## 3.0 Results

### 3.1 Participant characteristics

Of the 6,219 participants included in the present study, 55.4% were female, and the majority (55.3%) were aged 65-74 years with a mean age of 70 years. 93.7% of respondents classified themselves as being of white ethnic background, with 2.8% of Asian ethnic background, and only 0.7% of black African or Caribbean background. Approximately half of participants (48.6%) had a recorded height and weight, with a mean BMI of 25.3 kg/m^2^. The majority of respondents were married (62.2%), co-habiting (72.8%) and retired (69.5%). Most respondents did not smoke (96.9%), drank alcohol (82.6%) and felt they ate a healthy diet (80.3%). 18.0% of respondents were classified as pre-frail, with 0.5% as frail and 26.2% reported that they were shielding at the time of the survey (table 1).

### 3.2 Physical activity before and after social distancing measures

Mean (SD) PA for participants prior to lockdown was 3,519 (2867) MET minutes/week. There was a significant reduction in mean MET minutes following implementation of lockdown to 3,186 (2673) MET minutes/week (p<0.001; table 2 & figure 1). 3,167 (50.9%) participants decreased their activity during lockdown by a mean (SD) of 1,957 (2025) MET minutes/week, 534 (8.6%) maintained the same level of activity, and 2,518 (40.5%) increased activity by a mean (SD) of 1,636 (1775) MET minutes/week. Mean sitting time increased by 276 MET minutes/week after lockdown (2,680) compared to before (2,404) (table 2).

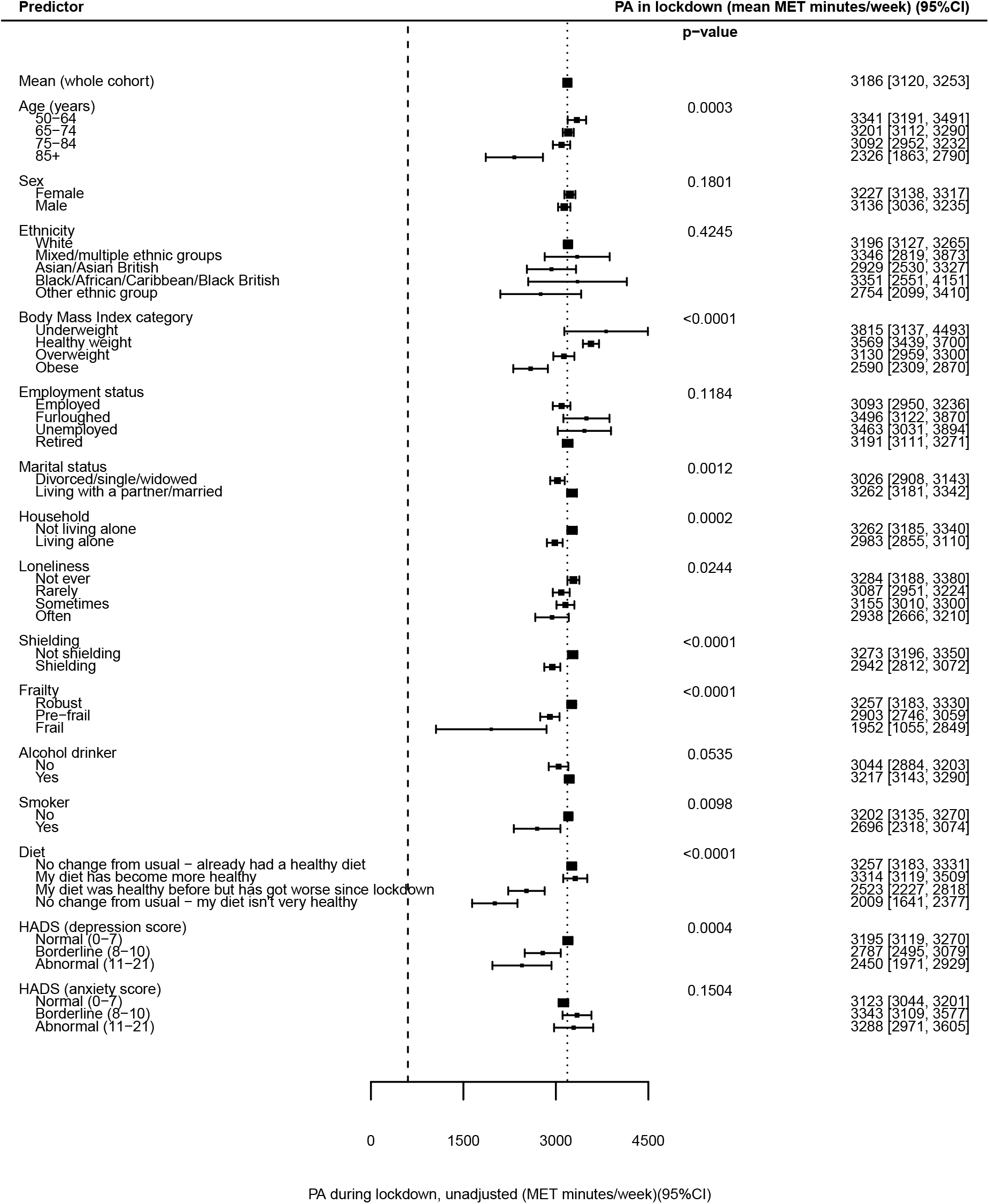

5,762 (92.7%) participants achieved at least the minimum guidance of 600 MET minutes/week of activity, as defined by WHO,^3^ prior to implementation of lockdown measures, slightly reducing to 5,672 (91.2%) following their introduction (p<0.001). 5,039 (81.0%) achieved 1,200 MET minutes/week before lockdown, with 4,904 (78.9%) achieving this after lockdown (p<0.001). Following lockdown, PA levels varied by month of survey completion, with the highest levels in June and lowest levels in July. There was no significant difference between self-reported PA before lockdown by month of survey completion (supplementary file 2: table 1 & figures 2-3).

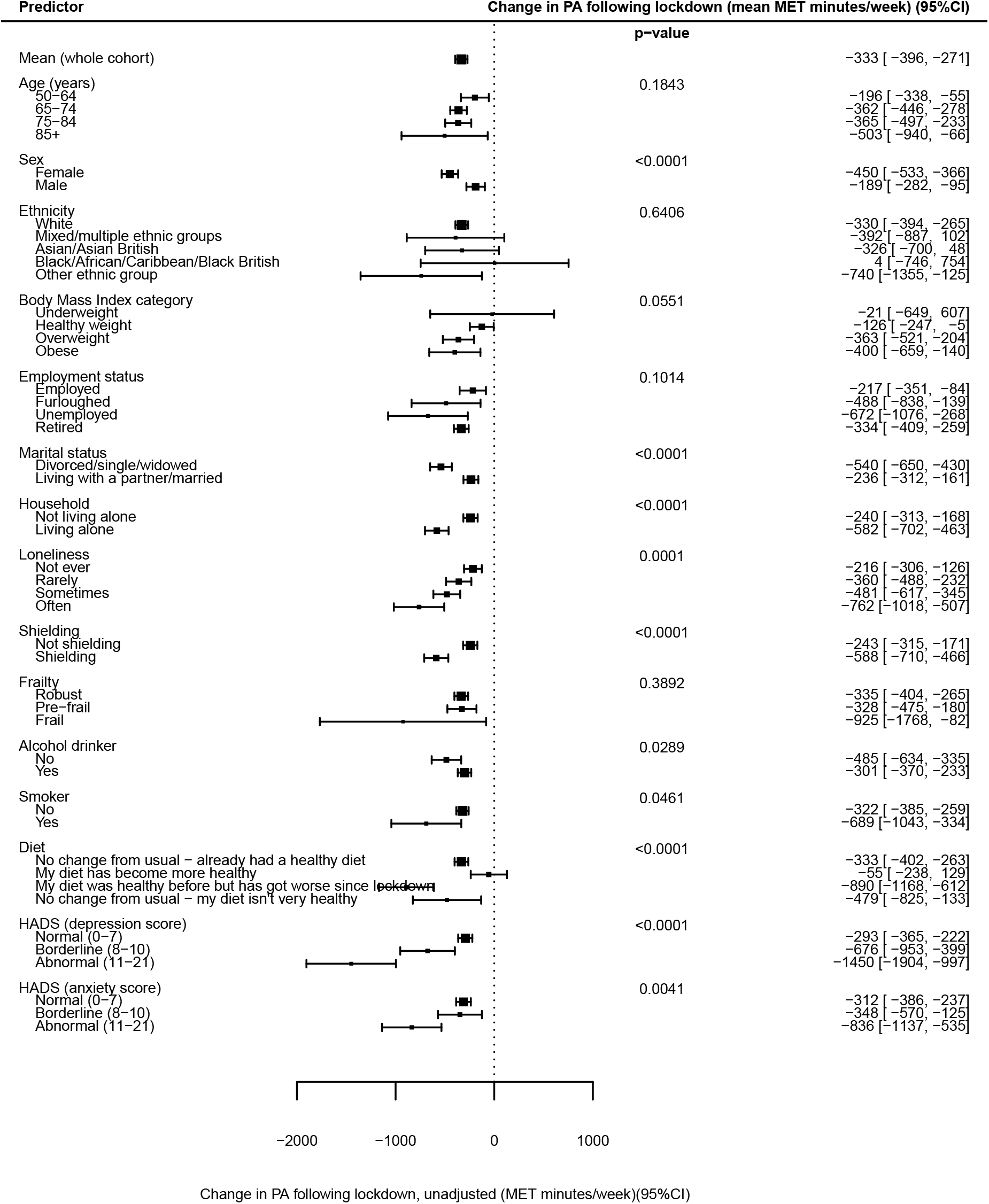

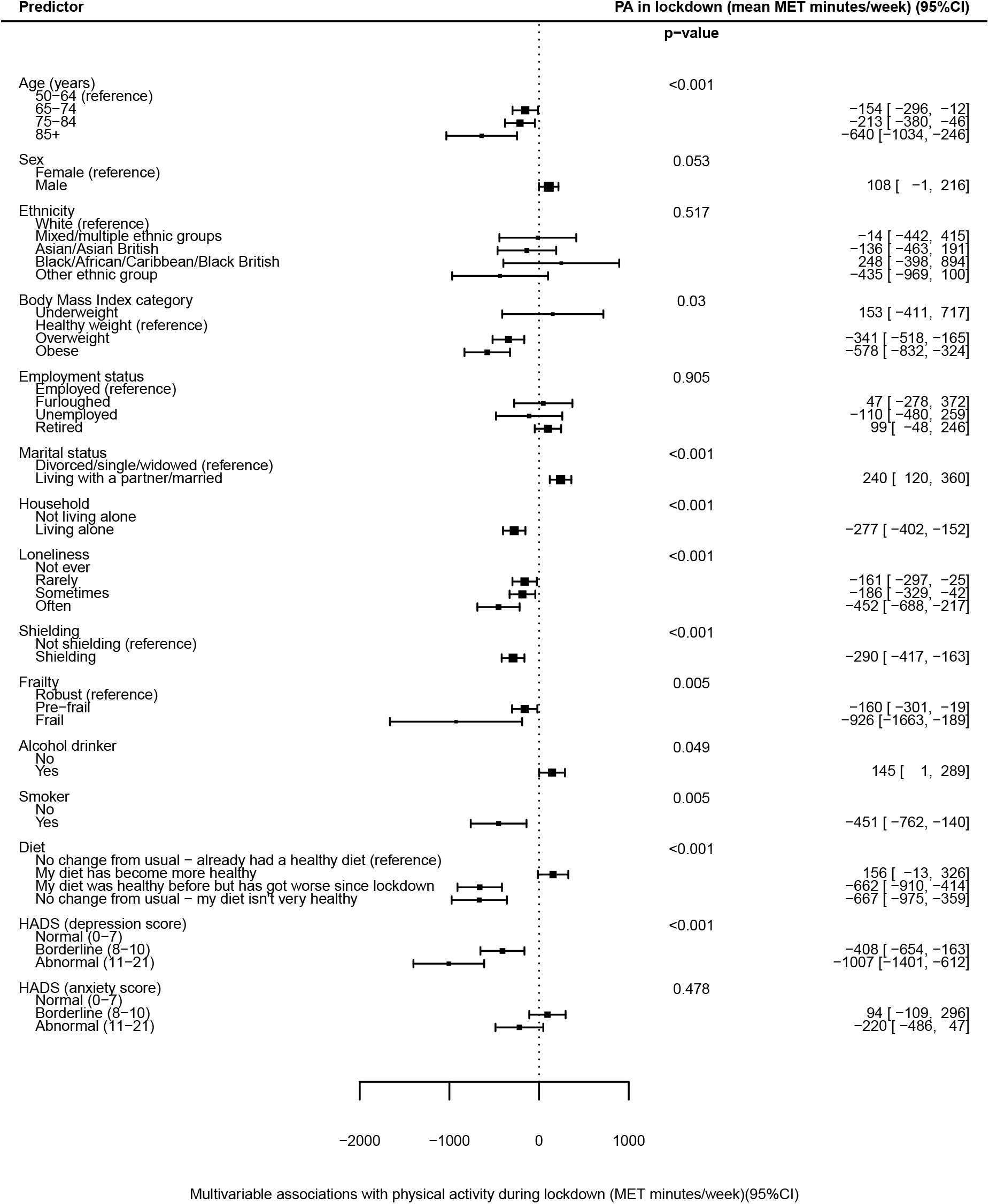

### 3.3 Predictors of physical activity after lockdown, and change from before lockdown

#### 3.3.1 Demographic and lifestyle factors

Univariable linear regression models showed statistically significant associations with lower PA after lockdown in older age groups but no evidence of differences in the change from before lockdown between age groups (p<0.001 and p=0.184, respectively; figures 1 & 2). After multivariable adjustment for sex, ethnicity, month of survey completion and pre-lockdown physical activity there was evidence of significantly lower levels of PA with increasing age, with adults aged 85 years and over doing on average 640 (95% CI: 246 to 1034) MET minutes/week less than those aged 50-64 years (figure 3). There was no significant difference in PA after lockdown in males and females (p=0.180), but females on average exhibited a greater decline in PA from before lockdown than males (450 vs 189 MET minutes/week less respectively; p<0.001; figures 1 & 2). After multivariable adjustment, including age, there was only a small and borderline significant difference in PA after lockdown between gender (PA in males on average 108 MET minutes/week more than females; 95% CI: -1 to 216; figure 3). No significant associations were seen between PA after lockdown or change in PA according to ethnicity or employment status, before or after adjustment.

Lower levels of PA after lockdown were seen with increasing BMI category, in current smokers and in those reporting an unhealthy or worsening diet before and after adjustment (figure 1). After adjustment, a dose-response relationship was evident between lower PA and increasing BMI (p=0.030), with obese individuals doing 578 (95% CI: 324 to 832) MET minutes/week less than those of a healthy weight (figure 3). The denominator included in analyses of BMI was significantly lower than for other models, as BMI was unrecorded for 51.4% of participants. Current alcohol consumption was weakly associated with increased levels of PA in both univariable and multivariable models, with current drinkers reporting 145 MET minutes/week more than non-drinkers after adjustment (95% CI: 1 to 289; figures 2 & 3).

#### 3.3.2 Associations with social isolation and loneliness

Participants who were divorced, single or widowed were, on average, less active after lockdown than those married or living with a partner (3,026 vs 3,262 MET minutes/week; p=0.001); and exhibited a greater decline in PA from before lockdown (540 vs 236 MET minutes/week less; p<0.001; figures 1 & 2). The association with PA after lockdown remained after adjustment, with those divorced, single or widowed doing on average 240 (95% CI: 120 to 360) MET minutes/week less (figure 3). Participants living alone were also less active than those co-habiting and showed greater reductions in PA from before lockdown. After adjustment for confounders and PA before lockdown, those living alone were doing 277 (95% CI: 152 to 402) MET minutes/week less than those co-habiting (figure 3).

Significant associations were seen between PA after lockdown and frequency of loneliness, with those ‘often’ experiencing loneliness achieving 2,938 MET minutes/week compared with 3,284 MET minutes/week in those ‘never’ experiencing loneliness (p=0.024; figure 1). Greater declines in PA from before lockdown were also seen with increasing loneliness (figure 2). After adjustment, PA after lockdown was significantly lower for those with increased frequency of loneliness (figure 3). After full adjustment including, in addition, household status, shielding status and frailty category, those experiencing loneliness ‘often’ reported 306 (95% CI: 60 to 552) MET minutes/week less activity than those ‘never’ lonely (supplementary file 2: table 4).

Significantly lower physical activity levels were recorded in those shielding and in participants categorised as pre-frail or frail (both p<0.001; figure 1). Larger declines in PA from before lockdown were seen in those shielding compared to those not shielding (588 vs 243 MET minutes/week less; p<0.001), but there was no significant difference in change in PA according to frailty category (p=0.389; figure 2). After adjustment, frail participants were doing 926 (95% CI: 189 to 1,663) MET minutes less on average than those classed as robust (figure 3). Participants who were shielding were doing an average of 290 (95% CI: 163 to 417) MET minutes/week less than those not shielding (figure 3).

#### 3.3.3 Associations with depression and anxiety

Symptoms of depression were associated with lower levels of PA during lockdown, with those meeting the criteria for depression reporting 2,450 MET minutes/week compared to 3,195 MET minutes/week in those with normal scores (p<0.001; figure 1). There was no strong association with anxiety scores. Mean change in PA from before lockdown was associated with both depression and, in contrast to absolute PA levels, with anxiety scores. Participants with depression reported 1,450 MET minutes/week less on average after lockdown compared with before, while those with normal scores reported 293 MET minutes/week less (p<0.001). Similarly, in those with anxiety, PA reduced by 836 MET minutes/week compared to 312 MET minutes/week in those with normal scores (p=0.004; figure 2).

After adjustment, those meeting the criteria for depression on the HADS scale had significantly lower PA levels than those with normal scores, doing on average 1,007 (95% CI: 1401 to 612) MET minutes/week less (figure 3). There remained no statistically significant association between anxiety score and physical activity after adjustment.

## 4.0 Discussion

### 4.1 Main findings

Data from the CCRR study show that participants experienced, on average, a significant decrease in PA after the introduction of lockdown in the UK when compared with before, together with an increase in sitting time. When adjusted for age, sex, ethnicity, month of survey completion and baseline physical activity, factors strongly associated with a reduction in PA include; increased age, increased BMI, frailty, current smoking, and a change to a less healthy diet. Factors associated with social isolation were also significantly associated with a reduction in PA: those divorced, single or widowed, living alone, shielding or reporting increased frequency of loneliness did significantly less PA after lockdown. Furthermore, a strong association was also seen with lower PA during lockdown in those with depression, but not for those with anxiety.

### 4.2 The effect of lockdown on physical activity

There was a reduction in PA in over half of our participants, and a decrease in mean levels of PA by 333 MET minutes/week following the introduction of lockdown measures in the UK. This was accompanied by an increase in sitting time by 276 minutes per week, an adverse finding given the adverse health impacts associated with increased sedentary and sitting time.^20^ These findings correlate with other studies from the UK (a decrease in 25% of adults aged over 20 years following lockdown),^21^ Spain^22^ and China,^23^ and from a global survey collected in 8 different languages,^24^ despite the differences in outdoor exercise permissions between countries. Reductions in PA may impact disproportionately across society. We found that increasing age associated with a reduction in PA after lockdown, corresponding with that seen in Japan, with a 26.5% (65 minutes) decrease in total physical activity in adults aged 65 to 84.^25^ A self-reported study in the UK found that those with a diagnosis of obesity, hypertension, lung disease, depression or a disability were more likely to reduce PA during lockdown.^21^

### 4.3 Social relationships, loneliness, and physical activity

Individuals for whom social engagement was more likely to be restricted, such as those who were shielding, divorced, single, widowed, or living alone, were more likely to have lower levels of PA after lockdown, and to have declined to a greater extent. Similarly, those who subjectively reported feeling lonely were more likely to have lower PA levels, and greater declines from before lockdown. These associations remained significant after multivariable adjustment.

Associations between health behaviours, including PA, and social relationships have been noted previously. Data from the English Longitudinal Study of Ageing (ELSA) showed that socially isolated respondents were less likely to report healthy diets, and more likely to smoke.^7^ Crucially, they showed reduced activity counts in socially isolated individuals (measured by accelerometer) in a sample of adults older than 50 years,^8^ and reduced self-reported moderate to vigorous physical activity.^7^ This is particularly important given that isolated and lonely individuals are at an increased risk of morbidity and mortality from cardiovascular events, with the majority of this association mediated by risk factors which include physical inactivity.^26^ Fixed effect models from the ELSA cohort show that social disengagement, domestic isolation and loneliness are associated with measures of poorer physical performance, and although they appear to be independent of physical activity, may still be associated along the causal pathway.^27^ Studies of spousal pairs found that both men and women in married couples had greater levels of PA than their single counterparts,^28^ and changes in PA are positively associated with changes in the PA of a spouse.^29^ Increasing PA is associated with larger,^30,31^ more diverse^32^ and more heterogenous (in terms of PA) social networks, and having more physically active people in a social network is associated with being more active.^33^

The interaction between social relationships and PA levels may be bi-directional. Levels of PA are influenced by multiple factors at different levels, including individual (psychological, genetic); interpersonal (social networks); environmental (social, built, natural); and regional or global determinants.^34^ Social networks might influence PA through social support for individuals to take up and maintain activity, but also by regulating social norms, and associating PA with social connections or attachments.^35^ There may also be increased opportunities for PA^33^ when social networks are present.

### 4.4 Mood, health behaviours and physical activity

In those reporting symptoms of depression, there were significantly lower levels of PA and a significant decrease in activity when compared to before lockdown. These findings correlate with those from the UK,^36^ Australia,^37^ and Spain,^38^ which found inverse associations between physical activity levels and poor mental health. Similarly, a cross sectional study of Brazilian adults who were self-isolating found lower odds of symptoms of anxiety or depression in those who were performing over 30 or 15 minutes per day of moderate or vigorous activity respectively, and higher odds in those with prolonged sedentary time over 10 hours.^39^ The associations between PA and mental health are well known, with positive impacts on wellbeing,^40^ and reduced incidence and severity of symptoms of mental ill-health.^41–43^ Therefore, these findings are unsurprising, although the interaction between PA and reduced markers of mental ill-health in older adults may be bi- directional. Moreover, social isolation and loneliness may mediate some of this effect: previous data from the CCRR cohort showed an interaction between social isolation, loneliness, and female gender with worsening depression and anxiety over lockdown.^44^ We found no statistically significant difference in PA during lockdown with anxiety symptoms, at odds with previous studies.^36^ However, the trajectory of anxiety symptoms is not known, and it is not clear whether anxiety symptoms pre-dated the introduction of lockdown.

### 4.5 Health behaviours and physical activity

A decrease in PA was associated with other detrimental health behaviours, including unhealthy diet and smoking. A similar tendency of clustering of unhealthy behaviours during the COVID-19 pandemic was noted in a cohort of patients in Spain with type 2 diabetes mellitus, who showed an increase in sugary foods and snack consumption alongside an increase in sitting time, and a decrease in time spent walking or doing moderate physical activity during lockdown when compared to beforehand.^45^ That detrimental health behaviours might coincide in response to lockdown shows the importance of targeted interventions for certain groups. Interestingly, alcohol consumption was seen to be a protective factor in our cohort, and this does not tie with other findings on the negative associations with increased alcohol use during the COVID-19 pandemic.^46^ This may be due to the specific demographic features of our cohort, but the possibility of alcohol consumption being associated with social interaction in this group cannot be excluded.

### 4.6 Limitations

This study has several limitations which may impact the generalisability of our findings. First, the CCRR cohort appear more physically active than the general population. 90% of participants in CCRR achieved minimum UK ^4^ and WHO ^3^ guidance, both before and following lockdown. Over 78% achieved double this amount, and mean levels of PA were at least five times greater than the minimum recommendation. In contrast, only 61% of UK adults aged 55-74 years achieve minimum recommended levels.^2^ Despite this, CCRR participants may still not be active enough for major health gains. A 2016 systematic review and meta-analysis suggested that optimal risk reduction for breast and colorectal cancer, diabetes, ischaemic heart disease and stroke events were obtained from physical activity at 3000-4000 MET minutes per week.^47^

Second, there are differences in demography between the CCRR cohort and the general population of the UK, which may explain the higher levels of PA we observed. 93% of CCRR respondents identify as white/Caucasian ethnicity. The Active Lives Survey demonstrated a difference in those achieving minimum activity levels in White British individuals (65%) and those from Black (58%) and Asian (54%) ethnicities.^2^ Third, the CCRR survey relies on self-report, using the short form IPAQ. IPAQ data is well validated across diverse participants up to the age of 65 years ^10^ and a study of the performance of the IPAQ in older Japanese adults demonstrated adequate validity.^48^ However, results from self-reporting tools for PA only weakly correlate with those from objective measures, such as accelerometers and pedometers.^49–52^ Finally, recall bias and seasonal changes in physical activity may also have impacted on the results. The CCRR survey was collected in April-July 2020, with participants asked to recall PA levels in the week before lockdown, which over time may become less reliable. However, no significant differences were found in the mean PA levels reported before lockdown according to month of survey completion and although there were apparent differences in PA during lockdown by month, we were able to adjust for this in multivariable models. The CCRR prospective cohort study is ongoing, with follow-up questionnaires sent to participants at regular intervals. When complete this will allow for long-term impacts to be measured, accounting for seasonal variation.

### 4.7 Conclusions

Findings from our CCRR study suggest a significant decline in average physical activity levels in older adults following the introduction of lockdown measures during the COVID-19 pandemic. Lower activity levels after lockdown were strongly linked to older age, and to those with objective markers of social isolation, subjective feelings of loneliness and symptoms of depression. Strategies and targeted interventions to increase and sustain PA levels in older adults are needed to mitigate the adverse health impacts not only of COVID- 19 related lockdowns, but of social isolation in general, and should consider social relationships in their design and implementation.

## 5.0 Summary boxes

### What is already known on this topic

- Physical inactivity adversely affects older adults: almost two-thirds of adults over 75 years old are not sufficiently physically active for good health
- Social isolation and loneliness are associated with increased morbidity and mortality, and decreased physical activity; lockdowns for Covid-19, although crucial, may exacerbate this

### What this study adds

- Physical activity decreased in older adults following implementation of lockdown measures in the UK
- Those with factors suggesting increased social isolation, loneliness and depression were particularly susceptible to lower levels of physical activity after lockdown
- Interventions designed to increase physical activity in older adults should take account of social relationships in their design and implementation, and there is a case for specific resources to help protect socially isolated individuals during pandemic-related lockdowns

## Data Availability

This is an ongoing study, but anonymised data can be provided upon request for the purposes of further data analysis, and can be requested from the Data Management Co-ordinator, Parthenia Giannakopoulou

## Transparency declaration

The lead authors confirm that the submitted manuscript is an honest, accurate and transparent account of the study being reported. No important aspects of the study have been omitted.

## Ethics approval

This research was approved by the Imperial College Research and Ethics Committee (ICREC) and Joint Research Compliance Office (22/04/2020; 20IC5942). All participants were required to provide informed consent before taking part in the study. Data collected as a part of this study are anonymized and kept strictly confidential in accordance with the UK General Data Protection Regulations (2016).

## Data sharing

This is an ongoing study, but anonymised data can be provided upon request for the purposes of further data analysis, and can be requested from the Data Management Co- ordinator, Parthenia Giannakopoulou

## Dissemination declaration

Participants in the CHARIOT cohort are informed by regular newsletter of all publications pertaining to the cohort.

## Acknowledgements

Work towards this article was in part supported by the National Institute for Health Research (NIHR) Applied Research Collaboration Northwest London and Imperial Biomedical Research Centre (BRC). DS and TB are supported by NIHR academic clinical fellowships. The views expressed in this publication are those of the authors and not necessarily those of the National Institute for Health Research or the Department of Health and Social Care. Imperial College London is the sponsor for the CCRR study, and has no influence on the direction or content of the work. There was no external financial funding for the study.

We are grateful to Lesley Williamson, Monica Munoz-Troncoso, Snehal Pandya and Emily Pickering (CHARIOT register and facilitator team); Mariam Jiwani, Rachel Veeravalli, Islam Saiful, Danielle Rose, Susie Gold, Rachel Nejade and Shehla Shamsuddin (Imperial College London student volunteers); Stefan McGinn-Summers, Neil Beckford, Inthushaa Indrakumar and Kristina Lakey (Departmental administrative staff in AGE); Dinithi Perera (departmental manager); Heather McLellan-Young (project manager); Helen Ward, James McKeand, Geraint Price, Josip Car, Christina Atchison, Nicholas Peters, Aldo Faisal, and Jennifer Quint (investigator team contributing to CCRR survey design, development and improvement).

## How patients were involved in the creation of this article

Older adult volunteers (60-80 years of age) from various social and cultural backgrounds provided feedback on the survey content. This feedback was incorporated into the survey design.

## Licence

The Corresponding Author has the right to grant on behalf of all authors and does grant on behalf of all authors, <u>a worldwide licence</u> to the Publishers and its licensees in perpetuity, in all forms, formats and media (whether known now or created in the future), to i) publish, reproduce, distribute, display and store the Contribution, ii) translate the Contribution into other languages, create adaptations, reprints, include within collections and create summaries, extracts and/or, abstracts of the Contribution, iii) create any other derivative work(s) based on the Contribution, iv) to exploit all subsidiary rights in the Contribution, v) the inclusion of electronic links from the Contribution to third party material where-ever it may be located; and, vi) licence any third party to do any or all of the above.

---

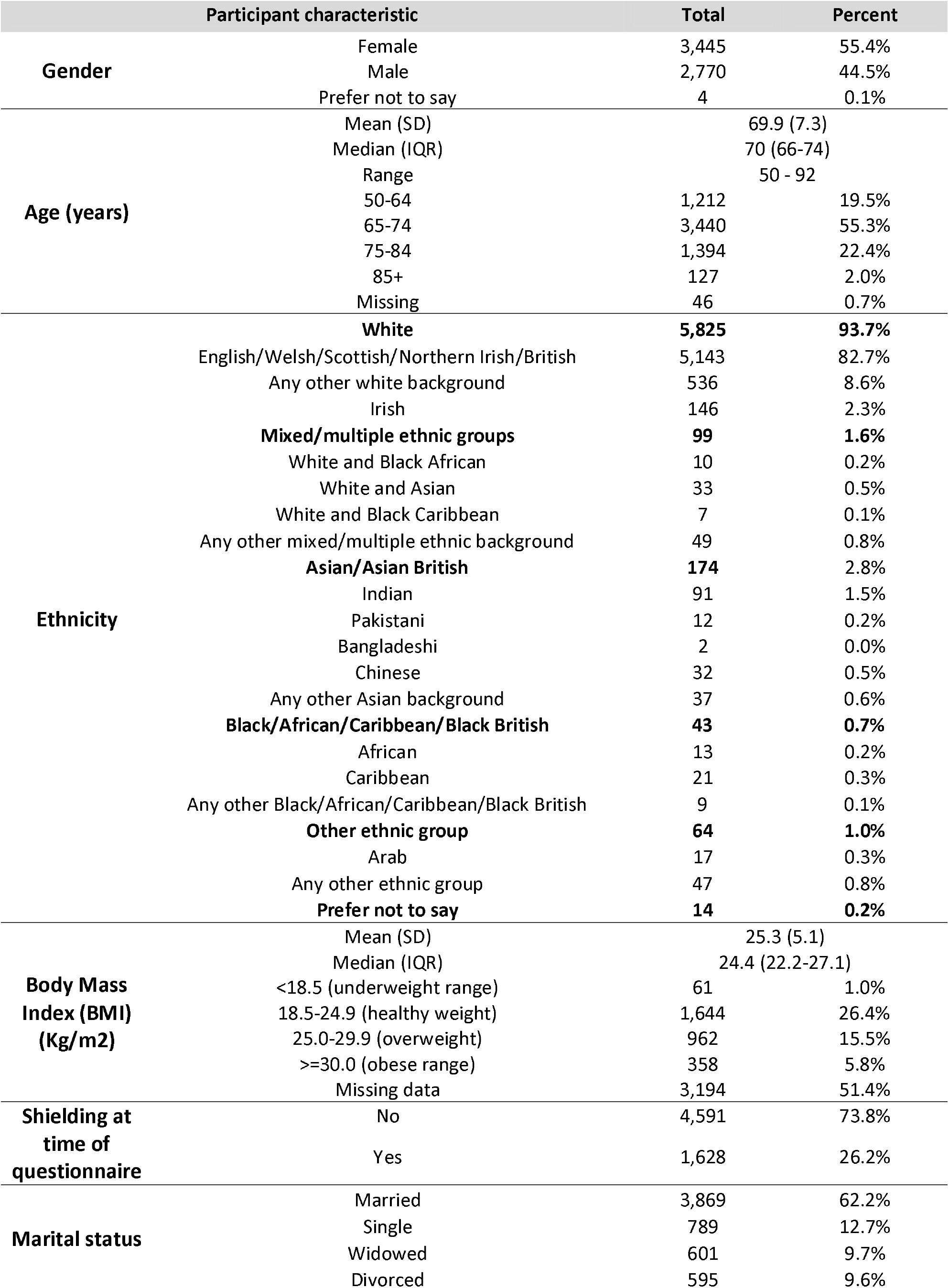
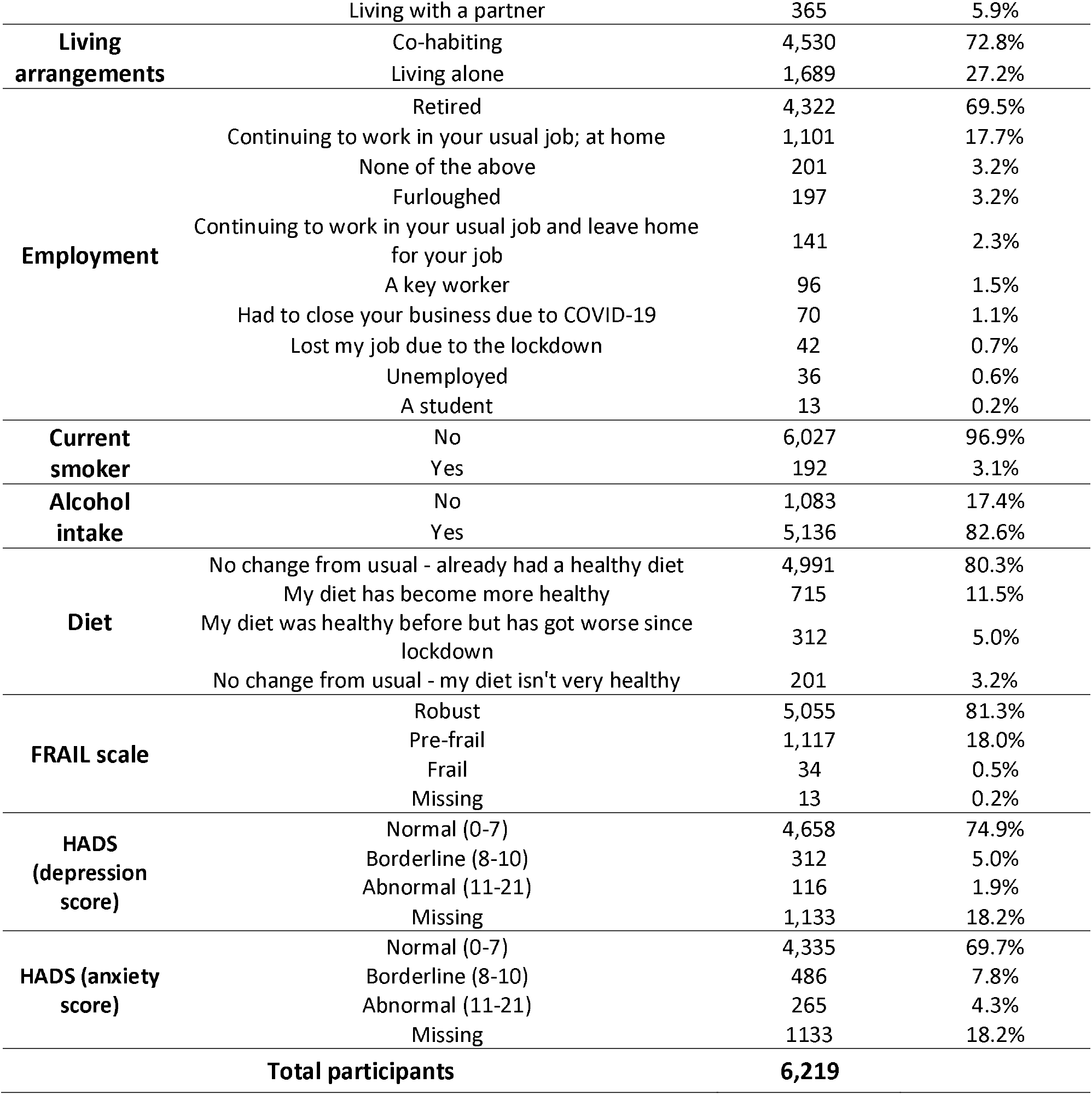

| Physical activity type |  | Before | During | p value for difference |
| --- | --- | --- | --- | --- |
| **Vigorous activity** | Mean (SD) minutes/week | 145 (276) | 135 (253) | 0.004 |
|  | Median (IQR) minutes/week | 40 (0 - 180) | 10 (0 - 180) |  |
| **Moderate activity (minutes/week)** | Mean (SD) minutes/week | 292 (430) | 245 (374) | <0.001 |
|  | Median (IQR) minutes/week | 120 (0 - 360) | 120 (0-360) |  |
| **Walking (minutes/week)** | Mean (SD) minutes/week | 462 (460) | 403 (408) | <0.001 |
|  | Median (IQR) minutes/week | 360 (150 - 630) | 315 (150 - 525) |  |
| **Sitting (minutes/week) \*** | Mean (SD) minutes/week | 2404 (1137) | 2680 (1181) | <0.001 |
|  | Median (IQR) minutes/week | 2100 (1680 - 2940) | 2520 (1680 - 3360) |  |
| **MET minutes/week** | Mean (SD) minutes/week | 3519 (2867) | 3185 (2673) | <0.001 |
|  | Median (IQR) minutes/week | 2772 (1386 - 4650) | 2440 (1386 - 4185) |  |

