## Supplementary material for "The impact of social restrictions during the COVID-19 pandemic on the physical activity levels of older adults: a baseline analysis of the CHARIOT COVID-19 Rapid Response prospective cohort study": caption

**Table and figure captions**

Table 1: Participant characteristics for 6,219 participants with complete data on physical activity; HADS – Hospital Anxiety and Depression Score

Table 2: Physical activity and sitting time for recipients before and following introduction of lockdown measures. Data presented as minutes per week with both mean (standard deviation) and median (interquartile range) shown. p-values from paired t-test; *denominator 6,023; MET - Metabolic Equivalent of Task

Figure 1: Forest plot of unadjusted univariable associations with physical activity (PA) during lockdown. Data presented as mean MET minutes/week +/- 95% confidence interval. Heavy dashed line – 600 MET minutes/week (WHO minimal physical activity guideline for adults); light dashed line – mean MET minutes for the whole cohort. See also supplementary file 2: table 2; HADS – Hospital Anxiety and Depression Score; MET – Metabolic Equivalent of Task; PA – Physical Activity; WHO – World Health Organization

Figure 2: Forest plot of unadjusted mean change in physical activity (PA) for all variables (mean MET minutes/week +/- 95% confidence interval). Negative values indicate a decline in activity after lockdown compared to before lockdown. See also supplementary file 2: table 2; HADS – Hospital Anxiety and Depression Score; MET – Metabolic Equivalent of Task; PA – Physical Activity

Figure 3: Forest plot of multivariable associations with physical activity after lockdown, adjusted for age, sex, ethnicity, month of year of survey completion and baseline physical activity. Data presented as mean MET minutes/week +/- 95% confidence interval, compared to the reference group, with negative values indicating lower physical activity than the reference. See also supplementary file 2: table 3; HADS – Hospital Anxiety and Depression Score; MET – Metabolic Equivalent of Task; PA – Physical Activity
