## Supplementary File 1 for "The impact of social restrictions during the COVID-19 pandemic on the physical activity levels of older adults: a baseline analysis of the CHARIOT COVID-19 Rapid Response prospective cohort study"

*Baseline Survey*

*Please answer all the questions in this survey before submitting it. Follow the*

*prompts for those questions that are not applicable to you.*

*Symptoms*

*Q1. In the last week, have you had a cough?*

*<1> No*

*<2> Yes*

*Q2. In the last week, have you experienced unusual shortness of breath (difficulty*

*breathing) compared to what's normal for you?*

*<1> No*

*<2> Yes, but it did not affect my normal activities*

*<3> Yes, it did affect my normal activities (eg walking short distances)*

*<4> Yes, even when I was sitting or lying down*

*Q3. In the last week, have you had a fever (feeling too hot) and did you take your*

*temperature?*

*<1> I have NOT felt feverish*

*<2> I have felt feverish but did not check my temperature*

*<3> I felt feverish and my temperature was equal to, or BELOW 38 degrees Celcius*

*<4> I felt feverish and my temperature measured ABOVE 38 degrees Celcius*

*Q4. In the last week, have you experienced any of these other symptoms? Please do*

*NOT include symptoms you experience on a regular basis due to a health condition*

*you already know about. Please tick all that apply:*

*<1> Loss of sense of smell*

*<2> Loss of sense of taste*

*<3> Decrease in appetite (skipping meals)*

*<4> Diarrhoea*

*<5> Nauseas and/or Vomiting*

*<6> Abdominal pain/tummy ache*

*<7> Chills (feeling too cold)*

*<8> Difficulty sleeping*

*<9> Felt more tired than normal*

*<10> Severe Fatigue*

*<11> Sneezing*

*<12> Chest pain / tightness*

*<13> Tightness in chest*

*<14> Sore throat*

*<15> Hoarse voice*

*<16> Runny nose*

*<17> Blocked nose*

*<18> Sore eyes*

*<19> Itchy eyes*

*<20> Headache*

*<21> Joint pain / aches*

*<22> Dizziness*

*<23> Muscle pain/aches*

*<99> None of these*

*If you answered, ‘None of these’, please skip Q5 and go to Q6.*

*Q5. Thinking about the 14 days before your symptoms started, had you been in*

*physical contact (within 2 metres / 6 feet) with someone who has a confirmed*

*diagnosis of coronavirus (Covid-19), or someone with the following symptoms: dry*

*cough, fever, loss of sense of smell, loss of sense of taste, shortness of breath or*

*difficulty breathing.*

*<1> Yes, and it was an individual within my household*

*<2> Yes, and it was an individual from outside my household*

*<3> No, not that I am aware of*

*QX Since COVID-19 emerged in January, but before the official lockdown started on*

*March 23rd 2020, which, if any of the following, have you experienced? Please do*

*NOT include symptoms you experience on a regular basis due to a health condition*

*you already know about. Please tick all that apply.*

*<1> New, continuous cough (coughing a lot for more than an hour, or have had 3*

*coughing episodes in 24 hours)*

*<2> High temperature (hot to touch on chest or back)*

*<3> Loss of sense of smell*

*<4> Loss of sense of taste*

*<5> Loss of appetite (skipping meals)*

*<6> Diarrhoea*

*<7> Vomiting*

*<8> Fatigue*

*<9> Sneezing*

*<10> Chest pain / tightness*

*<11> Sore throat*

*<12> Runny nose*

*<13> Itchy eyes*

*<14> Headache*

*<15> Joint pain / aches*

*<16> Muscle or joint pain*

*<99> None of these*

*If you answered, ‘None of these’, go to Q6.*

*QXa Approximately when did you start experiencing these symptoms?*

*[DD/MM/YYYY]*

*QXb Approximately how long did these symptoms last?*

*[Days: ]*

*QXX Thinking about the 14 days before your symptoms started, had you been in*

*physical contact (within 2 metres / 6 feet) with someone who has a confirmed*

*diagnosis of coronavirus (Covid-19), or someone with the following symptoms: dry*

*cough, fever, loss of sense of smell, loss of sense of taste, shortness of breath or*

*difficulty breathing.*

*<1> Yes, and it was an individual within my household*

*<2> Yes, and it was an individual from outside my household*

*<3> No, not that I am aware of*

*Q6 Now, thinking about the period prior to last week, but after the official lockdown*

*started on 23rd March 2020 , which, if any of the following, have you experienced?*

*Please do NOT include symptoms you experience on a regular basis due to a health*

*condition you already know about. Please tick all that apply.*

*<1> Fever (feeling too hot)*

*<2> New persistent cough*

*<3> Shortness of breath affecting normal activities*

*<4> Loss of sense of smell*

*<5> Loss of sense of taste*

*<6> Decrease in appetite (skipping meals)*

*<7> Diarrhoea*

*<8> Nauseas and/or vomiting*

*<9> Abdominal pain/tummy ache*

*<10> Chills (feeling too cold)*

*<11> Difficulty sleeping*

*<12> Felt more tired than normal*

*<13> Severe fatigue*

*<14> Sneezing*

*<15> Chest pain*

*<16> Tightness in chest*

*<17> Sore throat*

*<18> Hoarse throat*

*<19> Runny nose*

*<20> Blocked nose*

*<21> Sore eyes*

*<22> Itchy eyes*

*<23> Headache*

*<24> Dizziness*

*<25> Joint pain / aches*

*<26> Muscle pain/aches*

*If you answered, ‘None of these’, go to Q8.*

*Q6a Approximately when did you start experiencing these symptoms?*

*[DD/MM/YYYY]*

*Q6b Approximately how long did these symptoms last?*

*[Days: ]*

*Q7 Thinking about the 14 days before your symptoms started, had you been in*

*physical contact (within 2 metres / 6 feet) with someone who has a confirmed*

*diagnosis of coronavirus (Covid-19), or someone with the following symptoms: dry*

*cough, fever, loss of sense of smell, loss of sense of taste, shortness of breath or*

*difficulty breathing.*

*<1> Yes, and it was an individual within my household*

*<2> Yes, and it was an individual from outside my household*

*<3> No, not that I am aware of*

*Q8 Have you or anyone in your house been tested for coronavirus? Please tick all*

*that apply*

*<1> No testing*

*<2> I have not been tested -- BUT I think I have already had coronavirus and*

*recovered*

*<3> I was tested - positive result*

*<4> I was tested - awaiting result*

*<5> I was tested - negative result*

*<6> Household member tested - positive result*

*<7> Household member tested - awaiting result*

*<8> Household member tested - negative result*

*Q9 In the last week, has anyone in your household had a new cough or fever?*

*Not applicable*

*<1> No*

*<2> Yes*

*Q10 Have you had any healthcare contact since the lockdown started? Please tick*

*all that apply*

*<1> No*

*<2> Yes - remote appointment with my GP (phone/video)*

*<3> Yes - I attended my GP practice for an appointment*

*<4> Yes - remote appointment with hospital (phone/video)*

*<5> Yes - I attended hospital for an appointment*

*<6> Yes - attended Accident and Emergency*

*<7> Yes -- I was admitted to hospital (not because of coronavirus)*

*<8> Yes -- I was admitted to hospital with symptoms of coronavirus*

*<9> Yes – One or more remote calls to 111- home visit by ambulance*

*Q11. In the last week, have you been taking any medication for new symptoms?*

*<1> No*

*<2> Yes*

*<3> If yes, what medication?*

*Underlying conditions*

*For the following question, please remember that your answers are always treated*

*confidentially and are never analysed individually. We have provided you with a*

*"Prefer not to say" option if you would rather not share your experiences.*

*Q12 Which, if any, of the following chronic health conditions have you been*

*diagnosed with? (Please select all that apply. If you do not currently have a chronic*

*health condition, please select the 'None of these' option)*

*<1> Arthritis*

*<2> Asthma*

*<3> My doctor has told me I have severe asthma*

*<4> I am having cancer treatment*

*<5> Blood or bone marrow cancer, such as leukaemia*

*<6> Cystic fibrosis*

*<7> Chronic obstructive pulmonary disease (COPD)*

*<8> Diabetes*

*<9> Epilepsy*

*<10> Heart disease*

*<11> High blood pressure*

*<12> High cholesterol*

*<13> HIV/ AIDS*

*<14> Mental health condition*

*<15> Multiple Sclerosis*

*<16> I have had an organ transplant*

*<17> I have a condition that makes me much more likely to get infections*

*<18> I am taking medicine that weakens my immune system*

*<19> Dementia, Parkinson’s or other neurological disease*

*<98> Prefer not to say*

*<99> None of these*

*Contacts*

*[Q13 What is your date of birth:*

*Date ….. ]*

*Q14 What is your sex:*

*<1> Female*

*<2> Male*

*<3> Prefer not to say*

*Q15 What ethnic group best describes you? Please select one option only.*

*<1>*

*English / Welsh /*

*Scottish / Northern*

*Irish / British*

*<11>*

*Bangladeshi*

*<2>*

*Irish*

*<12>*

*Chinese*

*<3>*

*Gypsy or Irish*

*Traveller*

*<13>*

*Any other Asian*

*background*

*<4>*

*Any other White*

*background*

*<14>*

*African*

*<5>*

*White and Black*

*Caribbean*

*<15>*

*Caribbean*

*<6>*

*White and Black*

*African*

*<16>*

*Any other Black /*

*African /*

*Caribbean*

*background*

*<7>*

*White and Asian*

*<17>*

*Arab*

*<8>*

*Any other Mixed /*

*Multiple ethnic*

*background*

*<18 fixed>*

*Any other ethnic*

*group*

*<9>*

*Indian*

*<19 fixed>*

*Prefer not to say*

*<10>*

*Pakistani*

*Q16 Who else is CURRENTLY living in your household? Please tick all that apply*

*<1> I live by myself*

*<2> I live with my partner*

*<3> I live with my child/children aged under 18*

*<4> I live with my child/children aged over 18*

*<5> I live with family members other than partner / children*

*<6> I live with housemates*

*16a Where are you living during lockdown?*

*<1> My usual home*

*<2> not my usual home – keeping away from household members who are at high*

*risk coronavirus*

*<3> Not my usual home – other reason*

*Q17 How many people, including yourself, are there in your household? Please*

*include both adults and children. If you live alone, enter 1*

*Number*

*Q17a*

*For each household member ask age (in years) and sex*

*The following questions will ask you to report on how many people you have come*

*into contact with both inside and outside of your household.*

*A contact is defined as either:*

*• Direct skin-to-skin physical contact (e.g. kiss/embrace/handshake)*

*• Face-to-face conversation with another person which lasts over 3 mins, within*

*2m distance*

*• Being within 2m distance from another individual for over 5 mins*

*Note: if you contacted the same person in different times through the day, they*

*should be counted once.*

*Q18 How many different people did you have contact with, both inside your*

*household and while outside (after having left your household) in the past 7 days?*

*Enter 0 if you had no contacts in the last 7 days*

*1. (enter number)*

*2. Don’t know*

*Q19a Among the contacts you had, just from yesterday, both inside your household*

*and while outside (after having left your household), how many belonged to the*

*following age groups?*

*No contacts yesterday*

*0 to <10 years old (enter number)*

*10 to <20 years old (enter number)*

*20 to <30 years old (enter number)*

*30 to <40 years old (enter number)*

*40 to <50 years old (enter number)*

*50 to <60 years old (enter number)*

*60 to <70 years old (enter number)*

*70 to <80 years old (enter number)*

*80 to <90 years old (enter number)*

*90+ years old (enter number)*

*12. Don’t know*

*IF Q18 is NOT=0*

*Q19b How many different people did you come in contact with in the past 7 days*

*outside of your household?*

*Enter 0 if you had no contacts in the last 7 days outside of your household*

*1. (enter number)*

*2. Don’t know*

*IF Q19b is NOT=0*

*Q19c Among the contacts you had, just from yesterday, outside your household,*

*how many belonged to the following age groups?*

*No contacts yesterday*

*0 to <10 years old (enter number)*

*10 to <20 years old (enter number)*

*20 to <30 years old (enter number)*

*30 to <40 years old (enter number)*

*40 to <50 years old (enter number)*

*50 to <60 years old (enter number)*

*60 to <70 years old (enter number)*

*70 to <80 years old (enter number)*

*80 to <90 years old (enter number)*

*90+ years old (enter number)*

*Don’t know*

*IF Q19b is NOT=0*

*Q19c Among the contacts that you have had in the past 7 days outside your*

*household, how many contacts occurred at work?*

*Enter 0 if you had no contacts in the last 7 days outside of your household that*

*occurred at work*

*1. (enter number)*

*2. Don’t know*

*For the following questions please answer according to the following terms;*

*Self-isolation – refers to those who are symptomatic and self-isolating for 7*

*days from when symptoms started*

*Shielding – those in specific vulnerable groups staying at home for 12 weeks.*

*These groups would include those with underlying chronic health conditions:*

*cancers, respiratory disease, on immunosuppressants, those at increased risk*

*of infection or pregnant women with heart disease and/or those advised by the*

*NHS of their extremely vulnerable status’.*

*Household quarantine – 14-day quarantine period for all members of a*

*household from the first day of symptom onset in first case in that household*

*Social distancing and isolation*

*Q20 Are you currently in self-isolation?*

*<1> Yes*

*<2> No*

*If yes, for how long:…days*

*Q21 Are you currently shielding as per government guidelines for vulnerable groups?*

*<1> Yes*

*<2> No*

*Q22 Have you moved residence recently due to the pandemic? Y/N*

*Q23. Are you single, married, living with a partner, divorced, widowed?*

*Q24. Are you*

*<1> Continuing to work in your usual job; at home*

*<2> Continuing to work in your usual job and leave home for your job <3>*

*volunteering in response to the COVID pandemic*

*<4> a key worker*

*<5> unemployed*

*<6> retired*

*<7> furloughed (put on leave, still getting paid)*

*<8> had to close your business due to COVID-19*

*<9> lost my job due to the lockdown*

*<10> a student*

*<99> None of the above*

*Q25. How often are you now contacting friends/family members remotely*

*(Skype/Zoom/Mobile/landline phone etc)?*

*Several times per day, once a day, 2-3 x per week, 4-6 x per week, once a week,*

*less than once a week?*

*Q26 Overall, how are your relationships with other members of your household?*

*Not applicable*

*1 = worst*

*2*

*3*

*4*

*5*

*6*

*7*

*8*

*9*

*10 = best*

*Q27 If you are leaving your home, what activity is this for? Please tick all that apply*

*<1> I am not leaving my home*

*<2> Commute to work*

*<3> Essential shopping*

*<4> Exercise*

*<5> Other*

*Q28 Have you or anyone in your household received a letter or message informing*

*you that you are in the population at ‘high risk’ from coronavirus? Please tick all that*

*apply.*

*<1> No - Neither myself or anyone in my household is at 'high risk'*

*<2> No - but I think I should have*

*<3> No - but someone in my household is at 'high risk'*

*<4> Yes - letter about me*

*<5> Yes - letter about someone in my household*

*Health behaviours: dietary, alcohol and smoking*

*Q29 Do you drink alcohol?*

*<1> Yes (If yes trigger sub-questions)*

*Drinking less since lockdown*

*Drinking the same amount since lockdown*

*Drinking more since lockdown*

*How many units do you consume per week:…units*

*(half pint/ 300ml = approx. 1 unit, 175ml glass wine= approx. 2 units)*

*<2> No (If no, trigger sub-questions)*

*I never drink alcohol*

*I had already stopped drinking alcohol before lockdown*

*I stopped drinking alcohol when lockdown started*

*Q30 Do you smoke?*

*<1> Yes (if yes, trigger sub-questions)*

*Smoking less since lockdown*

*Smoking the same amount since lockdown*

*Smoking more since lockdown*

*If yes, how many cigarettes or roll-ups do you smoke per day:…*

*<2> No (if now, trigger sub-questions)*

*I never smoked*

*I had already stopped smoking before lockdown*

*I stopped smoking since the lockdown*

*Q30a) Has there been a change in your vaping (e-cigarettes) status since the*

*coronavirus lockdown?*

*<1> I never vaped*

*<2> I had already stopped vaping before*

*<3> I stopped vaping since the lockdown*

*<4> Vaping less*

*<5> Vaping the same amount*

*<6> Vaping more*

*Q31 Since the lockdown, are you managing to keep a healthy diet, for example,*

*eating fresh fruits and vegetables?*

*<1> No change from usual - already had a healthy diet*

*<2> No change from usual - my diet isn't very healthy*

*<3> My diet has become more healthy*

*<4> My diet was healthy before but has got worse since lockdown*

*Q32 On average, how many portions (or servings) of fruit and vegetables do you eat*

*per day?.......*

*- One portion is typically 80g, 3 heaped tablespoons of cooked veg or 1 cereal*

*bowl of mixed salad*

*- Three heaped tablespoons of beans and other pulse vegetables, such as*

*kidney beans, lentils and chickpeas, count as 1 portion.*

*- The following starchy vegetables should not be included – potatoes, yams,*

*cassava and plantain*

*Q32a Have you ever skipped meals due to difficulties accessing food as a result of*

*COVID-19?*

*Yes /No*

*If yes:*

*How many meals per week, on average have you missed?*

*<1> 1-3 meals per week*

*<2> 4-6 meals per week*

*<4> 7-9 meals per week*

*<5> 10 or more meals per week*

*Biometric data: height and weight*

*Q33 Please enter your weight: Kg*

*Q34 Please enter your height:…cm*

*Q35 Do you have a recent (from the past week) blood pressure?_____mm/Hg*

*Current Physical activity: International Physical Activity Questionnaire*

*We are interested in finding out about the kinds of physical activities that people do*

*as part of their everyday lives. The questions will ask you about the time you spent*

*being physically active in the last 7 days. Please answer each question even if you*

*do not consider yourself to be an active person. Please think about the activities you*

*do at work, as part of your house and garden work, to get from place to place, and in*

*your spare time for recreation, exercise or sport.*

*Think about all the vigorous activities that you did in the last 7 days. Vigorous*

*physical activities refer to activities that take hard physical effort and make you*

*breathe much harder than normal. Think only about those physical activities that you*

*did for at least 10 minutes at a time.*

*Q36: During the last 7 days, on how many days did you do vigorous physical*

*activities like heavy lifting, digging, aerobics, or fast bicycling?*

*_____ days per week*

*If no vigorous physical activities, skip to question 38*

*Q37: How much time did you usually spend doing vigorous physical activities on*

*one of those days? If you only exercised in hours or minutes, please input a ‘0’ in the*

*non-applicable field.*

*_____ hours per day*

*_____ minutes per day*

*Think about all the moderate activities that you did in the last 7 days. Moderate*

*activities refer to activities that take moderate physical effort and make you breathe*

*somewhat harder than normal. Think only about those physical activities that you*

*did for at least 10 minutes at a time.*

*Q38: During the last 7 days, on how many days did you do moderate physical*

*activities like carrying light loads or bicycling at a regular pace? Do not include*

*walking.*

*_____ days per week*

*If no moderate physical activities, skip to question 40*

*Q39: How much time did you usually spend doing moderate physical activities on*

*one of those days? If you only exercised in hours or minutes, please input a ‘0’ in the*

*non-applicable field.*

*_____ hours per day*

*_____ minutes per day*

*Think about the time you spent walking in the last 7 days. This includes at work*

*and at home, walking to travel from place to place, and any other walking that you*

*have done solely for recreation, sport, exercise, or leisure.*

*Q40: During the last 7 days, on how many days did you walk for at least 10 minutes*

*at a time?*

*_____ days per week*

*No walking, skip to question 42*

*Q41: How much time did you usually spend walking on one of those days? If you*

*only exercised in hours or minutes, please input a ‘0’ in the non-applicable field.*

*_____ hours per day*

*_____ minutes per day*

*The last question is about the time you spent sitting on weekdays during the last 7*

*days. Include time spent at work, at home, while doing course work and during*

*leisure time. This may include time spent sitting at a desk, reading, or sitting or lying*

*down to watch television.*

*Q42: During the last 7 days, how much time did you spend sitting on a week day?*

*If you only exercised in hours or minutes, please input a ‘0’ in the non-applicable*

*field.*

*_____ hours per day*

*_____ minutes per day*

*Previous Physical activity: International Physical Activity Questionnaire*

*These questions will ask you about the time you spent being physically active in the*

*7 days prior to implementation of social distancing measures (please use first*

*week of March 2020). Please answer each question even if you do not consider*

*yourself to be an active person. Please think about the activities you do at work, as*

*part of your house and garden work, to get from place to place, and in your spare*

*time for recreation, exercise or sport.*

*Think about all the vigorous activities that you did in the 7 days prior to social*

*distancing measures. Vigorous physical activities refer to activities that take hard*

*physical effort and make you breathe much harder than normal. Think only about*

*those physical activities that you did for at least 10 minutes at a time.*

*Q43: During the 7 days prior to social distancing measures (please use first week*

*of March 2020), on how many days did you do vigorous physical activities like*

*heavy lifting, digging, aerobics, or fast bicycling?*

*_____ days per week*

*If no vigorous physical activities, skip to question 45*

*Q44: How much time did you usually spend doing vigorous physical activities on*

*one of those days? If you only exercised in hours or minutes, please input a ‘0’ in the*

*non-applicable field.*

*_____ hours per day*

*_____ minutes per day*

*Think about all the moderate activities that you did in the 7 days prior to social*

*distancing measures. Moderate activities refer to activities that take moderate*

*physical effort and make you breathe somewhat harder than normal. Think only*

*about those physical activities that you did for at least 10 minutes at a time.*

*Q45: During the 7 days prior to social distancing measures (please use first week*

*of March 2020), on how many days did you do moderate physical activities like*

*carrying light loads or bicycling at a regular pace? Do not include walking.*

*_____ days per week*

*If no moderate physical activities, skip to question 47*

*Q46: How much time did you usually spend doing moderate physical activities on*

*one of those days? If you only exercised in hours or minutes, please input a ‘0’ in the*

*non-applicable field.*

*_____ hours per day*

*_____ minutes per day*

*Think about the time you spent walking in the 7 days prior to social distancing*

*measures. This includes at work and at home, walking to travel from place to place,*

*and any other walking that you have done solely for recreation, sport, exercise, or*

*leisure.*

*Q47: During the 7 days prior to social distancing measures (please use first week*

*of March 2020), on how many days did you walk for at least 10 minutes at a time?*

*_____ days per week*

*No walking, skip to question 49*

*Q48: How much time did you usually spend walking on one of those days? If you*

*only exercised in hours or minutes, please input a ‘0’ in the non-applicable field.*

*_____ hours per day*

*_____ minutes per day*

*The last question is about the time you spent sitting on weekdays during 7 days*

*prior to social distancing measures. Include time spent at work, at home, while*

*doing course work and during leisure time. This may include time spent sitting at a*

*desk, reading, or sitting or lying down to watch television.*

*Q49: During the 7 days prior to social distancing measures (please use first week*

*of March 2020), how much time did you spend sitting on a week day? If you only*

*exercised in hours or minutes, please input a ‘0’ in the non-applicable field.*

*_____ hours per day*

*_____ minutes per day*

*Frailty Questionnaire:*

*Q50: Are you fatigued?*

*<1> Yes*

*<2> No*

*Q51: Can you walk up one flight of stairs?*

*<1> Yes*

*<2> No*

*Q52: Can you walk around the block?*

*<1> Yes*

*<2> No*

*Q53: Do you have more than 5 illnesses?*

*<1> Yes*

*<2> No*

*Q54: Have you lost more than 5% of your weight in the past 6 months?*

*<1> Yes*

*<2> No*

*Qx Have you had a fall during the COVID lockdown period?*

*Yes/No*

*If yes:*

*What actions were taken (select multiple where applicable):*

*a) No follow-up required, I did not hurt myself*

*b) Pain medication*

*c) A hospital and/or GP appointment*

*d) A follow-up X-ray*

*e) Sling/plaster cast for a fracture*

*f) Surgery*

*Mood*

*a) Tick the box beside the reply that is closest to how you have been feeling*

*in the past week.*

*Don’t take too long over you replies: your immediate response is best.*

*Tick here*

*1.*

*Tick here*

*8.*

*I feel tense or 'wound up':*

*I feel as if I am slowed down:*

*Most of the time*

*Nearly all the time*

*A lot of the time*

*Very often*

*From time to time, occasionally*

*Sometimes*

*Not at all*

*Not at all*

*2.*

*9.*

*I still enjoy the things I used to*

*enjoy:*

*I get a sort of frightened feeling like*

*'butterflies' in the stomach:*

*Definitely as much*

*Not at all*

*Not quite so much*

*Occasionally*

*Only a little*

*Quite Often*

*Hardly at all*

*Very Often*

*3.*

*10.*

*I get a sort of frightened feeling as if*

*something awful is about to*

*happen:*

*I have lost interest in my appearance:*

*Very definitely and quite badly*

*Definitely*

*Yes, but not too badly*

*I don't take as much care as I should*

*A little, but it doesn't worry me*

*I may not take quite as much care*

*Not at all*

*I take just as much care as ever*

*4.*

*11.*

*I can laugh and see the funny side*

*of things:*

*I feel restless as I have to be on the*

*move:*

*As much as I always could*

*Very much indeed*

*Not quite so much now*

*Quite a lot*

*Definitely not so much now*

*Not very much*

*Not at all*

*Not at all*

*5.*

*12.*

*Worrying thoughts go through my*

*mind:*

*I look forward with enjoyment to*

*things:*

*A great deal of the time*

*As much as I ever did*

*A lot of the time*

*Rather less than I used to*

*From time to time, but not too often*

*Definitely less than I used to*

*Only occasionally*

*Hardly at all*

*6.*

*13.*

*I feel cheerful:*

*I get sudden feelings of panic:*

*Not at all*

*Very often indeed*

*Not often*

*Quite often*

*Sometimes*

*Not very often*

*Most of the time*

*Not at all*

*7.*

*14.*

*I can sit at ease and feel relaxed:*

*I can enjoy a good book or radio or TV*

*program:*

*Definitely*

*Often*

*Usually*

*Sometimes*

*Not Often*

*Not often*

*Not at all*

*Very seldom*

*Qx . I experience a general sense of emptiness*

*• Not ever*

*• Rarely*

*• Sometimes*

*• Often*

*Qy. There are plenty of people I can rely on when I have problems*

*• Not ever*

*• Rarely*

*• Sometimes*

*• Often*

*Qz. I miss having people around me*

*• Not ever*

*• Rarely*

*• Sometimes*

*• Often*

*Please check you have answered all the questions above.*

*b) For each of the 17 mood questions above, please also indicate if you are*

*feeling or experiencing this 1, less than; 2, the same as; or 3, more than before*

*social isolation was implemented.*

*<1> [1 or 2 or 3]*

*<2> [1 or 2 or 3]*

*<3> [1 or 2 or 3]*

*<4> [1 or 2 or 3]*

*<5> [1 or 2 or 3]*

*<6> [1 or 2 or 3]*

*<7> [1 or 2 or 3]*

*<8> [1 or 2 or 3]*

*<9> [1 or 2 or 3]*

*<10> [1 or 2 or 3]*

*<11> [1 or 2 or 3]*

*<12> [1 or 2 or 3]*

*<13> [1 or 2 or 3]*

*<14> [1 or 2 or 3]*

*<15> [1 or 2 or 3]*

*<16> [1 or 2 or 3]*

*<17> [1 or 2 or 3]*

*Q. People may have worries about Covid-19. Have you been worried about any of*

*the following and, if so, how much?*

*Not at all*

*Little*

*Some*

*Rather*

*much*

*Very*

*much*

*Getting Covid-19 infection and/or infecting*

*someone else*

*That a person close to me could get infected*

*with Covid-19*

*Being discriminated against or avoided*

*because of Covid-19*

*Impact of the Covid-19 epidemic on my own*

*economy and/or loss of my employment*

*Economic impact of the Covid-19 epidemic*

*on the global economy*

*The government's and/or health system’s*

*lack of ability to handle the Covid-19*

*pandemic situation, including the shortage of*

*food and other groceries*

*Imperial College Sleep Quality (ICSQ) Questionnaire*

*Instructions:*

*The following questions relate to your usual sleep habits for a period of one month*

*before and during a period of reduced social contact. Your answers should indicate*

*the most accurate reply for the majority of days and nights during these periods.*

*Please answer all questions.*

*1. During the period before reduced social contact, what time did you usually go to*

*bed at night: bed-time was ……………………………………………………*

*1b) During the period of reduced social contact, what time have you usually gone to*

*bed at night: bed-time is - ………………………………………………………......*

*2. During the period before reduced social contact, how long (in minutes) did it*

*usually take you to fall asleep each night: number of minutes - …………………*

*2b) During the period of reduced social contact, how long (in minutes) has it usually*

*taken you to fall asleep each night: number of minutes - ……………………………*

*3. During the period before reduced social contact, what time did you usually get*

*up in the morning: getting-up time was - ………………………………………*

*3b) During the period of reduced social contact, what time do you usually get up in*

*the morning: getting-up time is - ………………………………………………………….*

*4. During the period before reduced social contact, how many hours of actual*

*sleep did you get at night? (This may be different from the number of hours you*

*spent in bed): hours of sleep per night - …………………………………………*

*4b) During the period of reduced social contact, how many hours of actual sleep do*

*you get at night? (This may be different from the number of hours you spend in bed):*

*hours of sleep per night - ………………………………………………...*

*5. During the period before reduced social contact, how often did you have trouble*

*sleeping because you could not get to sleep within 30 minutes:*

*o Not ever*

*o Less than once a week*

*o Once or twice a week*

*o Three or more times a week*

*5b) During the period of reduced social contact, how often have you had trouble*

*sleeping because you could not get to sleep within 30 minutes:*

*o Not ever*

*o Less than once a week*

*o Once or twice a week*

*o Three or more times a week*

*6. During the period before reduced social contact, did you experience poor sleep*

*(restless and unable to sleep):*

*o Not ever*

*o Less than once a week*

*o Once or twice a week*

*o Three or more times a week*

*6b) During the period of reduced social contact, have you experienced poor sleep*

*(restless and unable to sleep):*

*o Not ever*

*o Less than once a week*

*o Once or twice a week*

*o Three or more times a week*

*7a) During the period before reduced social contact, did you experience loneliness*

*(felt isolated, with no companions):*

*o Not ever*

*o Rarely*

*o Sometimes*

*o Often*

*7b) During the period of reduced social contact, have you experienced loneliness*

*(felt isolated, with no companions):*

*o Not ever*

*o Rarely*

*o Sometimes*

*o Often*

*7c) During the period of reduced social contact, have you experienced loneliness: 1,*

*less than; 2, the same as; or 3, more than before social isolation was implemented*

*Select: [1 or 2 or 3]*

*Functional Activities Questionnaire*

*For each of the tasks below please rate your ability to carry out the task/activity*

*independently on the following scale:*

*1. I had no difficulty*

*2. I had some difficulty, but I completed the task/activity myself.*

*3. I need some assistance to complete the task/activity:*

*a) I did not need assistance prior to COVID-19 lockdown but need assistance*

*now to maintain social isolation/distancing*

*b) I could do the task/activity before the COVID-19 lockdown, but now would need*

*assistance even if it were not to maintain social distancing*

*c) I required assistance since before the COVID-19 lockdown*

*4. I needed others to do this for me,*

*a) I could do the task/activity myself or with assistance prior to COVID-19 lockdown*

*but need others to do it for me to maintain social isolation/distancing*

*b) I could do the task/activity myself or with assistance before the COVID-19*

*lockdown, but now would need others to do it for me even if it were not to maintain*

*social distancing*

*c) I required others to do it for me since before the COVID-19 lockdown*

*5. I am unsure if I require assistance (e.g., never did the task/activity or have not*

*done the task/activity over the past week)*

*Activities:*

*1. Writing cheques, paying bills, balancing cheque book, using an ATM cash*

*machine*

*Response: …………………*

*2. Assembling tax records, business affairs, or papers*

*Response: …………………*

*3. Shopping alone for household necessities, medicines or groceries*

*Response: …………………*

*4. Playing a game of skill, working on a hobby*

*Response: …………………*

*5. Heating water, making a cup of coffee, turning off stove after use*

*Response: …………………*

*6. Preparing a balanced meal*

*Response: …………………*

*7. Keeping track of current events*

*Response: …………………*

*8. Paying attention to, understanding, discussing TV, video, book, magazine*

*Response: …………………*

*9. Remembering appointments, family occasions, public holidays, to take*

*medications*

*Response: …………………*

*10. Travelling out of my neighbourhood by taxi, car, bus or train and making travel*

*arrangements.*

*Response: …………………*

*THANK YOU FOR COMPLETING THIS QUESTIONNAIRE. YOUR RESPONSES*

*HAVE BEEN SAVED AND SENT TO THE STUDY TEAM.*

*NHS health advice and information regarding the novel coronavirus can be*

*found here: https://www.nhs.uk/conditions/coronavirus-covid-19/*

*For Advice on Mental health we suggest using these links:*

*1. The NHS Every Mind Matters website has information on how to look after*

*your mental wellbeing while in isolation: https://www.nhs.uk/oneyou/every-mind-*

*matters/*

*2. The charity Mental Health UK have advice on managing mental health during*

*the coronavirus outbreak: https://mentalhealth-uk.org/help-and-information/covid-19-*

*and-your-mental-health/*

*3. The NHS recommends a range of mobile apps to help with mental wellbeing,*

*many of which are free to download: https://www.nhs.uk/apps-*

*library/category/mental-health/*

*4. If you need someone to talk to about your mental health, the charity*

*Samaritans have a helpline available 24 hours a day, 7 days a week:*

*a. Call: 116 123*

*b. or visit: https://www.samaritans.org/how-we-can-help/contact-samaritan/*

*For Advice on Physical activity we suggest using these links:*

*1. The NHS Live Well website has a range of free advice and programmes from*

*light activity to more strenuous exercises for those aged under 65:*

*https://www.nhs.uk/live-well/exercise/*

*2. The NHS Live Well website has a range of free advice and programmes from*

*light activity to more strenuous exercises for those aged 65 or older:*

*https://www.nhs.uk/live-well/exercise/physical-activity-guidelines-older-adults/*

*3. Tips, advice and guidance from Sport England on how to keep or get active in*

*and around your home: https://www.sportengland.org/stayinworkout*

*4. Stay Active at Home: a simple set of exercises designed for older people to*

*stay active at home: https://www.csp.org.uk/public-patient/keeping-active-and-*

*healthy/staying-healthy-you-age/staying-strong-you-age/strength*

*For Advice on Sleep we suggest using these links:*

*1. The NHS ten top tips to improve sleep: https://www.nhs.uk/live-well/sleep-and-*

*tiredness/10-tips-to-beat-insomnia/*

*2. The NHS recommends a range of mobile apps to help with sleep:*

*https://www.nhs.uk/apps-library/category/sleep/*

Supplementary Table 1: CCRR survey
