## Supplementary File 2 for "The impact of social restrictions during the COVID-19 pandemic on the physical activity levels of older adults: a baseline analysis of the CHARIOT COVID-19 Rapid Response prospective cohort study"

**Supplementary methods**

Metabolic Equivalent of Task (MET) calculation

Briefly, 1 MET equates to an individual’s resting energy expenditure. According to the IPAQ scoring protocol, 3.3 METS is considered equivalent to walking, and moderate and vigorous activity to be 4 and 8 METS, respectively. To calculate the continuous variable of total MET minutes a week, the self-reported duration (minutes) and frequency (days) of each of these PA categories is multiplied by the by the specified metric.

**Supplementary figures and tables**

Figure 1: Box-plot of distribution of MET minutes per week before and during lockdown for 6,219 participants with completed IPAQ data

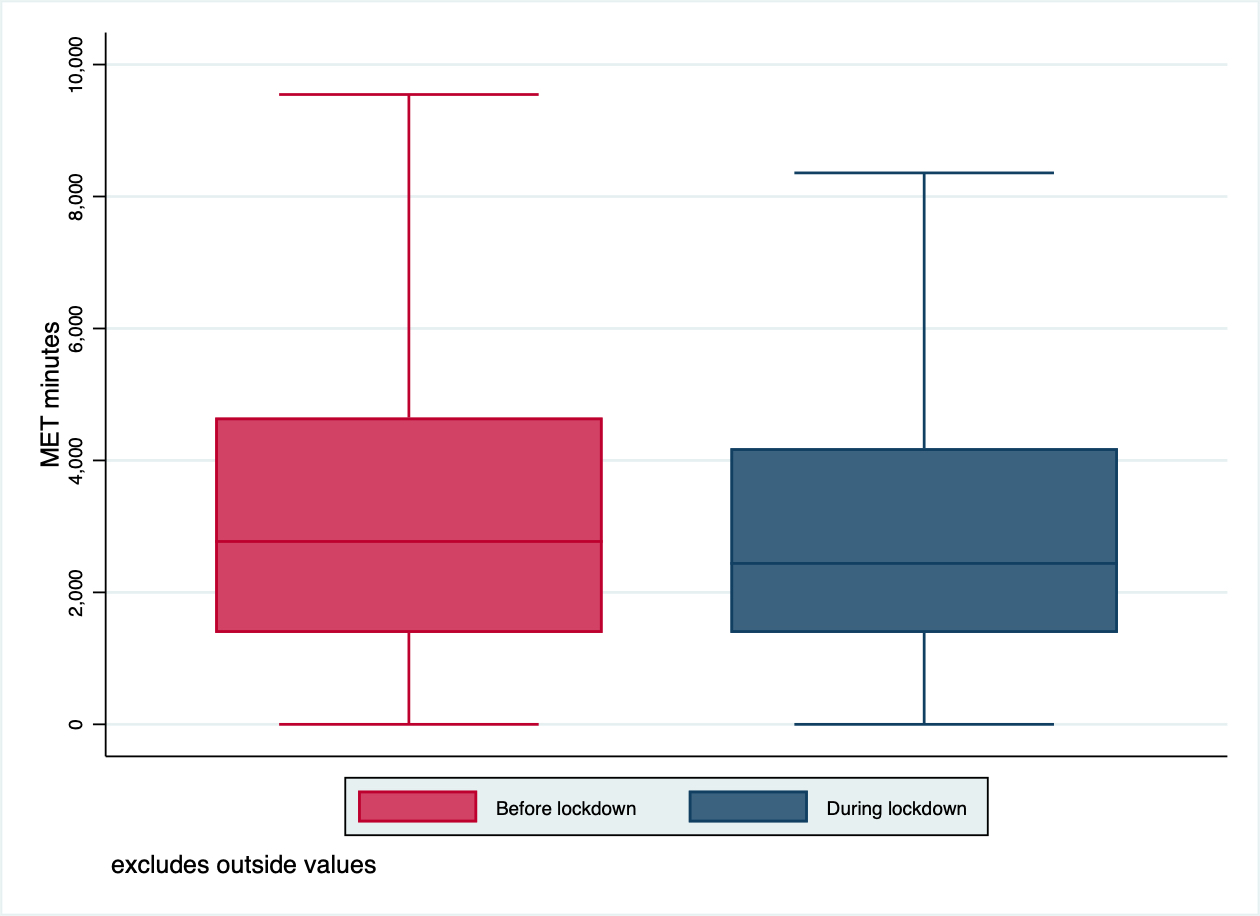

Figure 2: Box-plot of distribution of MET minutes per week after introduction of lockdown by month of survey completion for 6,219 participants with completed IPAQ data

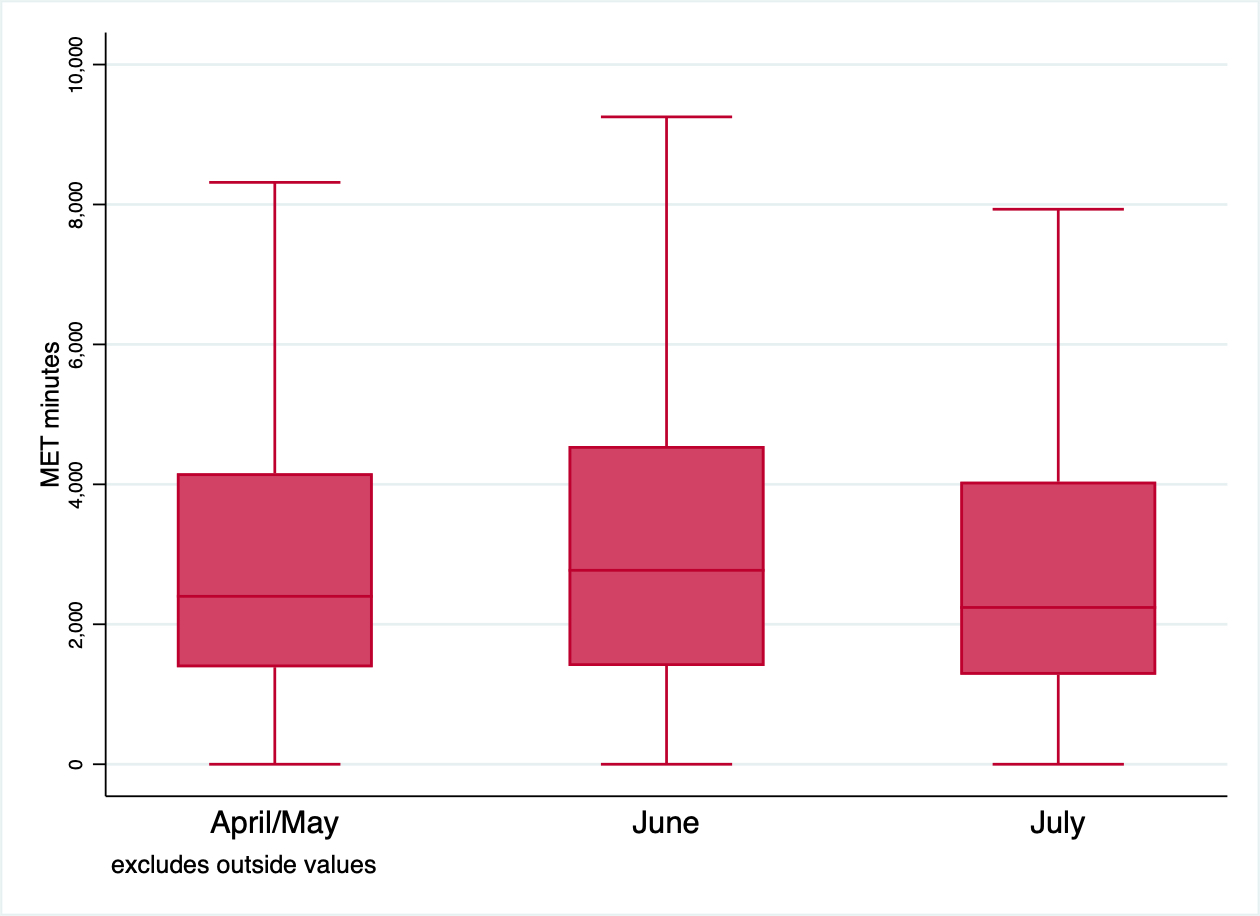

Table 1: Mean MET minutes after introduction of lockdown measures by month of survey completion

| **Month** | **Total** | **Percent** | **Mean MET minutes** | **p value^¶^** |
| --- | --- | --- | --- | --- |
| April/May* | 4975 | 80.0% | 3139 |  |
| June | 994 | 16.0% | 3470 | 0.0007 |
| July | 250 | 4.0% | 2967 |  |

* April (110) and May (4865) combined due to small numbers completed in April

**^¶^** p-value from linear regression models of MET minutes as dependent variable, against survey completion month as explanatory variable.

Figure 3: Box-plot of distribution of MET minutes per week before introduction of lockdown by month of survey completion for 6,219 participants with completed IPAQ data

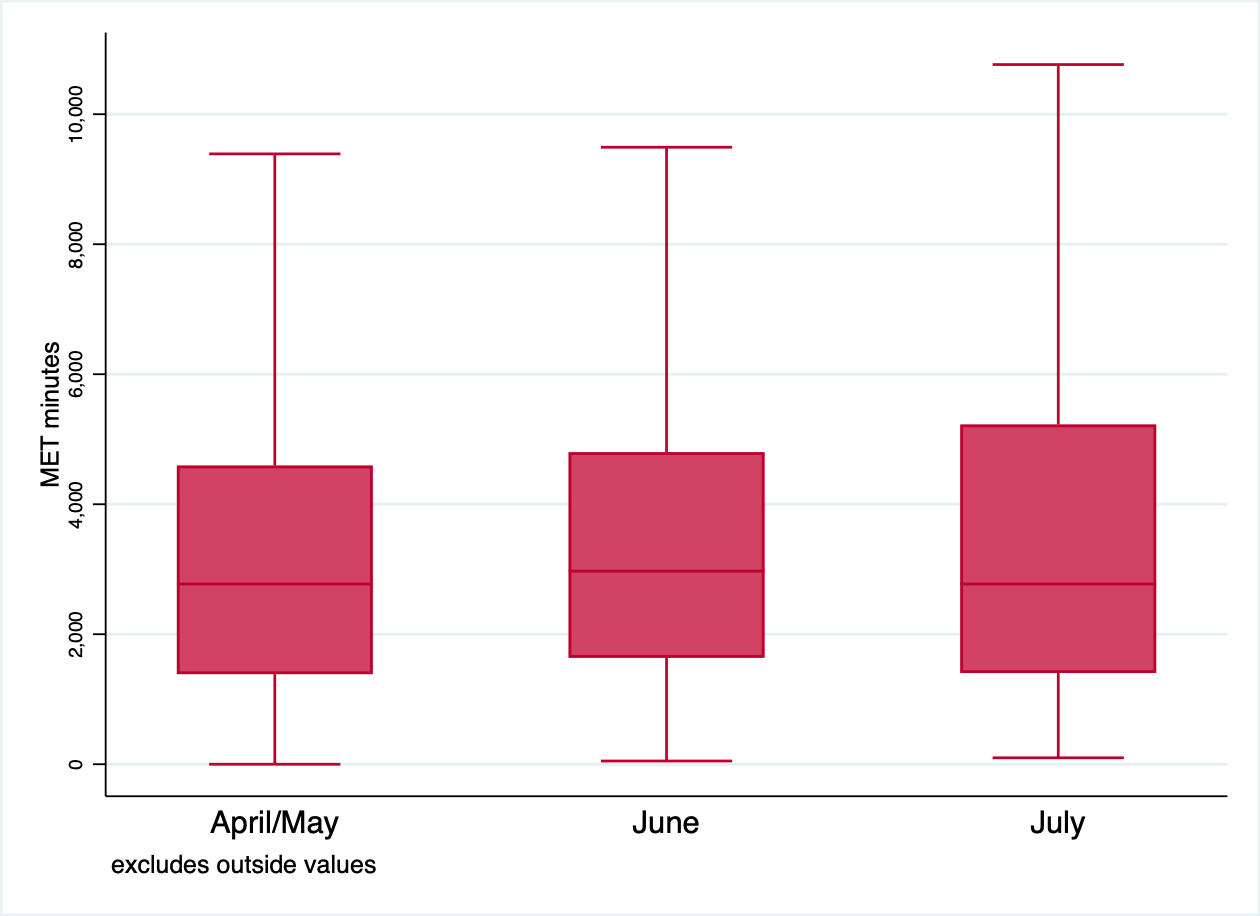

Linear regression models of MET minutes as dependent variable, against survey completion month as explanatory variable showed no significant association (p=0.1112).

Figure 4: Causal diagram representing factors impacting on change in physical activity after lockdown

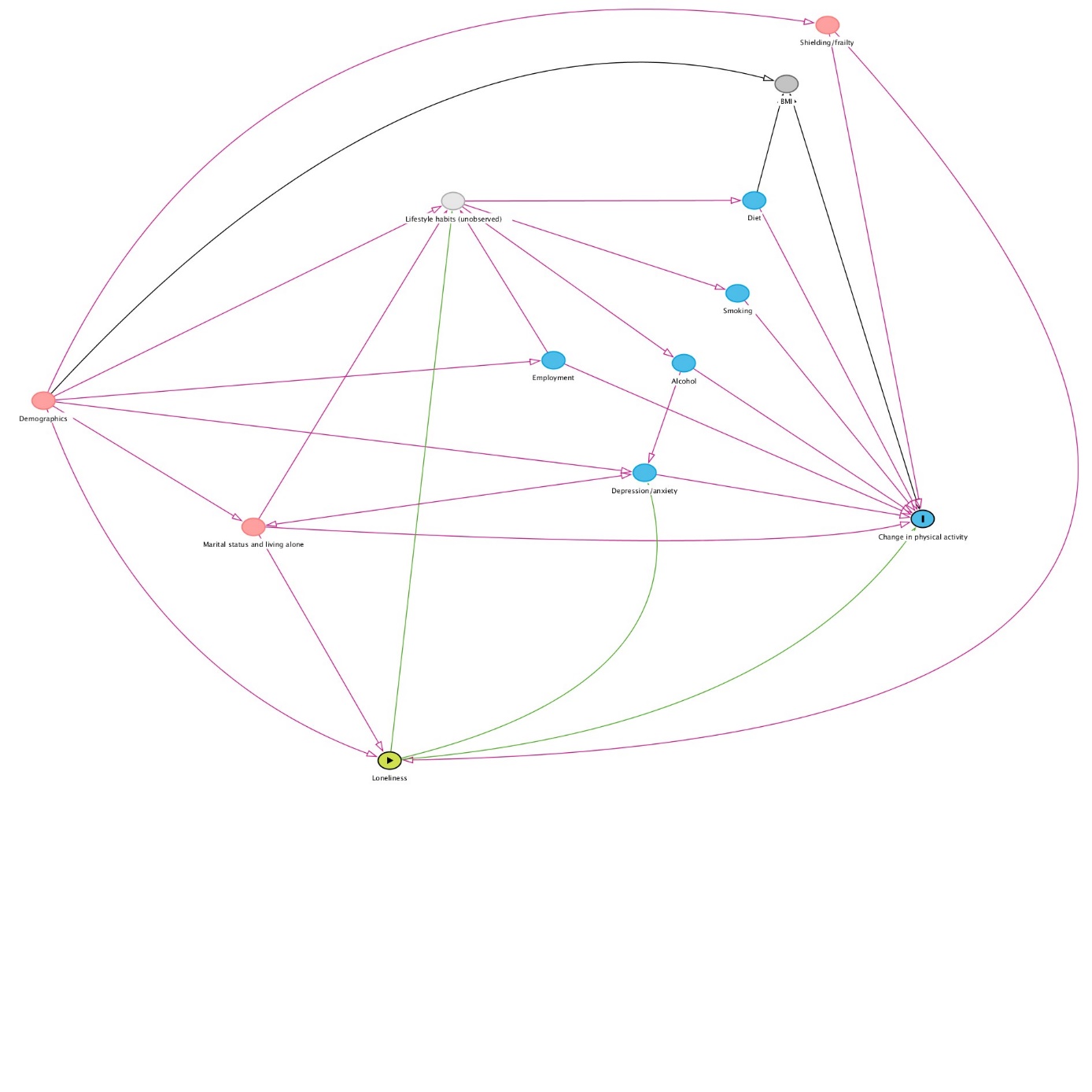

Table 2: Unadjusted associations in physical activity (MET minutes per week) after introduction of lockdown measures and change from before lockdown, from linear regression models. Note: negative values for change in activity indicate reduction after lockdown

|  | Physical activity after lockdown (MET minutes/week) | | | | Change in physical activity from before lockdown (MET minutes/week) | | | |
| --- | --- | --- | --- | --- | --- | --- | --- | --- |
| **Predictor** | **Mean** | **95% confidence interval** | | **p value** | **Mean** | **95% confidence interval** | | **p value** |
|  |  | **Lower** | **Upper** |  |  | **Lower** | **Upper** |  |
| **Mean (whole cohort)** | 3186 | 3120 | 3253 | - | -333 | -396 | -271 | - |
| **Age (years)** |  |  |  | <0.001 |  |  |  | 0.184 |
| 50-64 | 3341 | 3191 | 3491 |  | -196 | -338 | -55 |  |
| 65-74 | 3201 | 3112 | 3290 |  | -362 | -446 | -278 |  |
| 75-84 | 3092 | 2952 | 3232 |  | -365 | -497 | -233 |  |
| 85+ | 2326 | 1863 | 2790 |  | -503 | -940 | -66 |  |
| **Sex** |  |  |  | 0.180 |  |  |  | <0.001 |
| Female | 3227 | 3138 | 3317 |  | -450 | -533 | -366 |  |
| Male | 3136 | 3036 | 3235 |  | -189 | -282 | -95 |  |
| **Ethnicity** |  |  |  | 0.425 |  |  |  | 0.641 |
| White | 3196 | 3127 | 3265 |  | -330 | -394 | -265 |  |
| Mixed/multiple ethnic groups | 3346 | 2819 | 3873 |  | -392 | -887 | 102 |  |
| Asian/Asian British | 2929 | 2530 | 3327 |  | -326 | -700 | 48 |  |
| Black/African/Caribbean/Black British | 3351 | 2551 | 4151 |  | 4 | -746 | 754 |  |
| Other ethnic group | 2754 | 2099 | 3410 |  | -740 | -1355 | -125 |  |
| **Body Mass Index category** |  |  |  | <0.001 |  |  |  | 0.055 |
| Underweight | 3815 | 3137 | 4493 |  | -21 | -649 | 607 |  |
| Healthy weight | 3569 | 3439 | 3700 |  | -126 | -247 | -5 |  |
| Overweight | 3130 | 2959 | 3300 |  | -363 | -521 | -204 |  |
| Obese | 2590 | 2309 | 2870 |  | -400 | -659 | -140 |  |
| **Employment status** |  |  |  | 0.118 |  |  |  | 0.101 |
| Employed | 3093 | 2950 | 3236 |  | -217 | -351 | -84 |  |
| Furloughed | 3496 | 3122 | 3870 |  | -488 | -838 | -139 |  |
| Unemployed | 3463 | 3031 | 3894 |  | -672 | -1076 | -268 |  |
| Retired | 3191 | 3111 | 3271 |  | -334 | -409 | -259 |  |
| **Marital status** |  |  |  | 0.001 |  |  |  | <0.001 |
| Divorced/single/widowed | 3026 | 2908 | 3143 |  | -540 | -650 | -430 |  |
| Living with a partner/married | 3262 | 3181 | 3342 |  | -236 | -312 | -161 |  |
| **Household** |  |  |  | <0.001 |  |  |  | <0.001 |
| Not living alone | 3262 | 3185 | 3340 |  | -240 | -313 | -168 |  |
| Living alone | 2983 | 2855 | 3110 |  | -582 | -702 | -463 |  |
| **Loneliness** |  |  |  | 0.024 |  |  |  | <0.001 |
| Not ever | 3284 | 3188 | 3380 |  | -216 | -306 | -126 |  |
| Rarely | 3087 | 2951 | 3224 |  | -360 | -488 | -232 |  |
| Sometimes | 3155 | 3010 | 3300 |  | -481 | -617 | -345 |  |
| Often | 2938 | 2666 | 3210 |  | -762 | -1018 | -507 |  |
| **Shielding** |  |  |  | <0.001 |  |  |  | <0.001 |
| Not shielding | 3273 | 3196 | 3350 |  | -243 | -315 | -171 |  |
| Shielding | 2942 | 2812 | 3072 |  | -588 | -710 | -466 |  |
| **Frailty** |  |  |  | <0.001 |  |  |  | 0.389 |
| Robust | 3257 | 3183 | 3330 |  | -335 | -404 | -265 |  |
| Pre-frail | 2903 | 2746 | 3059 |  | -328 | -475 | -180 |  |
| Frail | 1952 | 1055 | 2849 |  | -925 | -1768 | -82 |  |
| **Alcohol drinker** |  |  |  | 0.054 |  |  |  | 0.029 |
| No | 3044 | 2884 | 3203 |  | -485 | -634 | -335 |  |
| Yes | 3217 | 3143 | 3290 |  | -301 | -370 | -233 |  |
| **Smoker** |  |  |  | 0.010 |  |  |  | 0.046 |
| No | 3202 | 3135 | 3270 |  | -322 | -385 | -259 |  |
| Yes | 2696 | 2318 | 3074 |  | -689 | -1043 | -334 |  |
| **Diet** |  |  |  | <0.001 |  |  |  | <0.001 |
| No change from usual - already had a healthy diet | 3257 | 3183 | 3331 |  | -333 | -402 | -263 |  |
| My diet has become more healthy | 3314 | 3119 | 3509 |  | -55 | -238 | 129 |  |
| My diet was healthy before but has got worse since lockdown | 2523 | 2227 | 2818 |  | -890 | -1168 | -612 |  |
| No change from usual - my diet isn't very healthy | 2009 | 1641 | 2377 |  | -479 | -825 | -133 |  |
| **HADS (depression score)** |  |  |  | <0.001 |  |  |  | <0.001 |
| Normal (0-7) | 3195 | 3119 | 3270 |  | -293 | -365 | -222 |  |
| Borderline (8-10) | 2787 | 2495 | 3079 |  | -676 | -953 | -399 |  |
| Abnormal (11-21) | 2450 | 1971 | 2929 |  | -1450 | -1904 | -997 |  |
| **HADS (anxiety score)** |  |  |  | 0.150 |  |  |  | 0.004 |
| Normal (0-7) | 3123 | 3044 | 3201 |  | -312 | -386 | -237 |  |
| Borderline (8-10) | 3343 | 3109 | 3577 |  | -348 | -570 | -125 |  |
| Abnormal (11-21) | 3288 | 2971 | 3605 |  | -836 | -1137 | -535 |  |

*HADS – Hospital Anxiety and Depression Score

Table 3: Results of multivariable linear regression models of physical activity after lockdown, adjusted for age, sex, ethnicity, month of survey completion and baseline physical activity. Data presented as mean MET minutes/week +/- 95% confidence interval compared to the reference group, with negative values indicating lower physical activity than the reference.

| **Predictor** | **Physical activity after lockdown (MET minutes/week)** | **95% confidence interval** | | **p value** | **Number of observations** |
| --- | --- | --- | --- | --- | --- |
|  |  | **Lower** | **Upper** |  |  |
| **Age (years)** |  |  |  | <0.001 | 6155 |
| 50-64 (reference) | - | - | - |  |  |
| 65-74 | -154 | -296 | -12 |  |  |
| 75-84 | -213 | -380 | -46 |  |  |
| 85+ | -640 | -1034 | -246 |  |  |
| **Sex** |  |  |  | 0.053 | 6155 |
| Female (reference) | - | - | - |  |  |
| Male | 108 | -1 | 216 |  |  |
| **Ethnicity** |  |  |  | 0.517 | 6155 |
| White (reference) | - | - | - |  |  |
| Mixed/multiple ethnic groups | -14 | -442 | 415 |  |  |
| Asian/Asian British | -136 | -463 | 191 |  |  |
| Black/African/Caribbean/Black British | 248 | -398 | 894 |  |  |
| Other ethnic group | -435 | -969 | 100 |  |  |
| **Body Mass Index category** |  |  |  | 0.030 | 2987 |
| Underweight | 153 | -411 | 717 |  |  |
| Healthy weight (reference) | - | - | - |  |  |
| Overweight | -341 | -518 | -165 |  |  |
| Obese | -578 | -832 | -324 |  |  |
| **Employment status** |  |  |  | 0.905 | 5958 |
| Employed (reference) | - | - | - |  |  |
| Furloughed | 47 | -278 | 372 |  |  |
| Unemployed | -110 | -480 | 259 |  |  |
| Retired | 99 | -48 | 246 |  |  |
| **Marital status** |  |  |  | <0.001 | 6155 |
| Divorced/single/widowed (reference) | - | - | - |  |  |
| Living with a partner/married | 240 | 120 | 360 |  |  |
| **Household** |  |  |  | <0.001 | 6155 |
| Not living alone (reference) | - | - | - |  |  |
| Living alone | -277 | -402 | -152 |  |  |
| **Loneliness** |  |  |  | <0.001 | 6077 |
| Not ever (reference) | - | - | - |  |  |
| Rarely | -161 | -297 | -25 |  |  |
| Sometimes | -186 | -329 | -42 |  |  |
| Often | -452 | -688 | -217 |  |  |
| **Shielding** |  |  |  | <0.001 | 6155 |
| Not shielding (reference) | - | - | - |  |  |
| Shielding | -290 | -417 | -163 |  |  |
| **Frailty** |  |  |  | 0.005 | 6142 |
| Robust (reference) | - | - | - |  |  |
| Pre-frail | -160 | -301 | -19 |  |  |
| Frail | -926 | -1663 | -189 |  |  |
| **Alcohol drinker** |  |  |  | 0.049 | 6155 |
| No (reference) | - | - | - |  |  |
| Yes | 145 | 1 | 289 |  |  |
| **Smoker** |  |  |  | 0.005 | 6155 |
| No (reference) |  |  |  |  |  |
| Yes | -451 | -762 | -140 |  |  |
| **Diet** |  |  |  | <0.001 | 6155 |
| No change from usual - already had a healthy diet (reference) | - | - | - |  |  |
| My diet has become more healthy | 156 | -13 | 326 |  |  |
| My diet was healthy before but has got worse since lockdown | -662 | -910 | -414 |  |  |
| No change from usual - my diet isn't very healthy | -667 | -975 | -359 |  |  |
| **HADS (depression score)** |  |  |  | <0.001 | 5038 |
| Normal (0-7) (reference) | - | - | - |  |  |
| Borderline (8-10) | -408 | -654 | -163 |  |  |
| Abnormal (11-21) | -1007 | -1401 | -612 |  |  |
| **HADS (anxiety score)** |  |  |  | 0.478 | 5038 |
| Normal (0-7) (reference) | - | - | - |  |  |
| Borderline (8-10) | 94 | -109 | 296 |  |  |
| Abnormal (11-21) | -220 | -486 | 47 |  |  |

HADS – Hospital Anxiety and Depression Score

Table 4: Multivariable linear regression model for physical activity after lockdown with loneliness, adjusted for age, sex, ethnicity, month of survey completion, baseline physical activity, living alone, marital status, shielding and frailty

| **Predictor** | **Physical activity after lockdown (MET minutes/week)** | **95% confidence interval** | | **p value** | **Number of observations** |
| --- | --- | --- | --- | --- | --- |
|  |  | **Lower** | **Upper** |  |  |
| **Loneliness** |  |  |  | 0.007 | 6077 |
| Not ever (reference) | - | - | - |  |  |
| Rarely | -127 | -265 | 11 |  |  |
| Sometimes | -107 | -256 | 42 |  |  |
| Often | -306 | -552 | -60 |  |  |
| Adjusted: age, sex, ethnicity, month of survey completion, baseline physical activity, living alone, marital status, shielding, frailty | | | | | |
